# Machine Learning-Driven Correction of Handgrip Strength: A Novel Biomarker for Neurological and Health Outcomes in the UK Biobank

**DOI:** 10.1101/2024.11.27.24318057

**Authors:** Kimia Nazarzadeh, Simon B. Eickhoff, Georgios Antonopoulos, Lukas Hensel, Caroline Tscherpel, Vera Komeyer, Federico Raimondo, Veronika Müller, Christian Grefkes, Kaustubh R. Patil

## Abstract

**Background:** Handgrip strength (HGS) is a significant biomarker for overall health, offering a simple, cost-effective method for assessing muscle function. Lower HGS is linked to higher mortality, functional decline, cognitive impairments, and chronic diseases. Considering the influence of anthropometrics and demographics on HGS, this study aims to develop a corrected HGS score using machine learning (ML) models to enhance its utility in understanding brain health and disease.

**Methods:** Using UK Biobank data, sex-specific ML models were developed to predict HGS based on three anthropometric variables and age. A novel biomarker, Δ*HGS*, was introduced as the difference between true HGS (i.e., directly measured HGS) and bias-free predicted HGS. The neural basis of true HGS and Δ*HGS* was investigated by correlating them to regional gray matter volume (GMV). Statistical analyses were performed to test their sensitivity to longitudinal changes in stroke and major depressive disorder (MDD) patients compared to matched healthy controls (HC).

**Results:** HGS could be accurately predicted using anthropometric and demographic features, with linear support vector machine (SVM) demonstrating high accuracy. Compared to true HGS, Δ*HGS* showed high reassessment reliability and stronger, widespread associations with GMV, especially in motor-related regions. Longitudinal analysis revealed that neither HGS nor Δ*HGS* effectively differentiated patients from matched HC at post time-point.

**Conclusion:** The proposed Δ*HGS* score exhibited stronger correlations with GMV compared to true HGS, suggesting it better represents the relationship between muscle strength and brain structure. While not effective in differentiating patients from HC at post time-point, the increase in Δ*HGS* from pre to post time-points in patient cohorts may indicate improved utility for monitoring disease progression, treatment efficacy, or rehabilitation effects, warranting further longitudinal validation.

## Background

Handgrip strength (HGS) is broadly recognized as a reliable and non-invasive biomarker of overall health. It is typically measured isometrically using a hydraulic hand dynamometer, an instrument that shows good reassessment reliability [1,2]. This measurement involves participants squeezing the dynamometer with maximum effort without any hand or arm movement, thus measuring the isometric grip force. Direct measured HGS offers several advantages, including low cost, ease of administration, and a strong predictive value for various health outcomes [1,3–6]. Beyond, HGS is related to the grey matter volume (GMV) of the brain, which in turn has been used as a marker for neuropathological changes in neurodegenerative and psychiatric diseases. GMV has also been associated with protective factors such as muscular strength [1]. Lower GMV is associated with lower HGS [7]. On the other hand, stronger HGS is associated with higher GMV in a wide array of brain regions like the ventral striatum, hippocampus, thalamus, pallidum, putamen, brain stem, temporal pole, and parahippocampal gyrus [1]. Through its associations with physical capabilities and with structural brain integrity, HGS offers insights into the neurobiological mechanisms related to muscle strength [1]. This relationship, however, is influenced by genetics, physical fitness, mental health as well as anthropometrics, and demographic characteristics, emphasizing the need for a comprehensive examination of these associations [1].

HGS serves as a marker of overall health because it can be affected through different mechanisms. Lower HGS in older adults is associated with adverse health outcomes, such as increased mortality risk, diminished functional mobility, cognitive impairments, and a range of health issues, including metabolic diseases like diabetes, and neurological conditions including stroke and major depressive disorder (MDD) [8–10]. Moreover, reduced HGS is not only linked to higher disease recovery times [11] but also functions as a valuable biomarker for assessing recovery and prognosis in stroke patients, with evidence linking a decrease in HGS after stroke (post-stroke) compared to before stroke (pre-stroke) levels [12,13]. These attributes have established HGS as an indicator of muscle strength and general health in clinical settings [1,8,11,14]. While understanding longitudinal changes in HGS in patients can help risk assessment and improve monitoring, to date it remains understudied due to lack of longitudinal data.

Anthropometric factors such as height, body mass index (BMI), and waist-to-hip ratio (WHR) are directly linked to HGS [15–20]. For instance, BMI and height correlate positively with greater strength [17,19]. Conversely, WHR, which reflects the proportion of abdominal fat relative to hip circumference, often exhibits an inverse relationship with HGS. Greater abdominal fat relative to hip size is frequently associated with lower HGS [18]. Age is another critical factor affecting HGS. Generally, HGS increases during childhood and adolescence, peaks in early adulthood, and then declines with higher age, particularly after 40. This decline is often more pronounced in older adults, especially those over 75, where the rate of decrease accelerates [16]. The aging process results in progressive muscle loss, further impacting HGS [16], [21]. Sex differences also contribute significantly to variations in HGS [22,23], influencing both baseline strength levels and decline patterns. Males and females exhibit distinct patterns in disease progression and anthropometric measurements, necessitating sex-specific considerations in clinical and research settings [24]. The interpretation of HGS is thus most meaningful when normalized to anthropometric and demographic factors, rather than relying on raw measured values. Relative HGS, which accounts for these variables, may offer a more precise assessment of an individual’s neuromuscular deficit and its relationship with brain structure and diseases, potentially improving its utility as a health indicator [8,11,12,25]. In particular, a refined HGS interpretation can open up new possibilities for early disease detection, monitoring treatment efficacy, guiding recovery processes, and predicting long-term health outcomes across various conditions.

Machine learning (ML) based predictive modelling offers individual-level predictions, establishing ML as a transformative tool in modern clinical practice [20,26]. ML can be used to predict HGS using anthropometric and demographic features. This predicted HGS captures the variance explained by the features and thus can be used to develop a relative HGS score. In this study, we tested the hypothesis that the difference between true HGS and predicted HGS can serve as a biomarker for muscular strength relating to the structural integrity of the brain and disease-related changes. Using data from the UK Biobank (UKB), we developed sex-specific ML models to predict HGS using anthropometrics and demographic variables in healthy individuals. We applied statistical bias-correction techniques to enhance prediction accuracy and introduced a novel score called Δ*HGS*, defined as the difference between the true HGS and bias-free predicted HGS. We then investigated the brain basis Δ*HGS* based on correlation with regional GMV followed by the investigation of its sensitivity to longitudinal changes in patient cohorts from two diseases known to influence HGS: stroke and MDD. The key innovation lies in our longitudinal design, capturing HGS scores at two time points: before (pre) and after (post) disease onset. This approach enabled us to examine how Δ*HGS* and true HGS change over time in patients compared to healthy controls (HC) groups.

## Methods

### Participants and data

We used data from the UK Biobank database with more than half a million adult participants recruited from a total of 22 assessment centers across the United Kingdom (UK) with baseline assessment between 2006 and 2010. The baseline assessment included a wide range of demographic data, physical measurements, clinical and health-related information, and the completion of a touchscreen questionnaire [27,28]. A subset of participants was invited back in 2012-2013 for the first repeat assessment. During this repeat visit, additional data were collected, although no brain imaging data were collected at either the baseline or the first repeat assessment. Starting in 2014, a subsample was invited to assessment centers for brain imaging, with follow-up imaging assessments beginning in 2019. In this study, participants were categorized into three groups: healthy controls (HC) and two patient cohorts: stroke and MDD. Group definitions were established using inclusion and exclusion criteria based on the International Classification of Diseases, 10th revision (ICD-10) coding system. We required that all anthropometrics, demographics, and HGS be complete for each participant in the final study sample to ensure no missing values (Fig. 1).

**Fig. 1.**
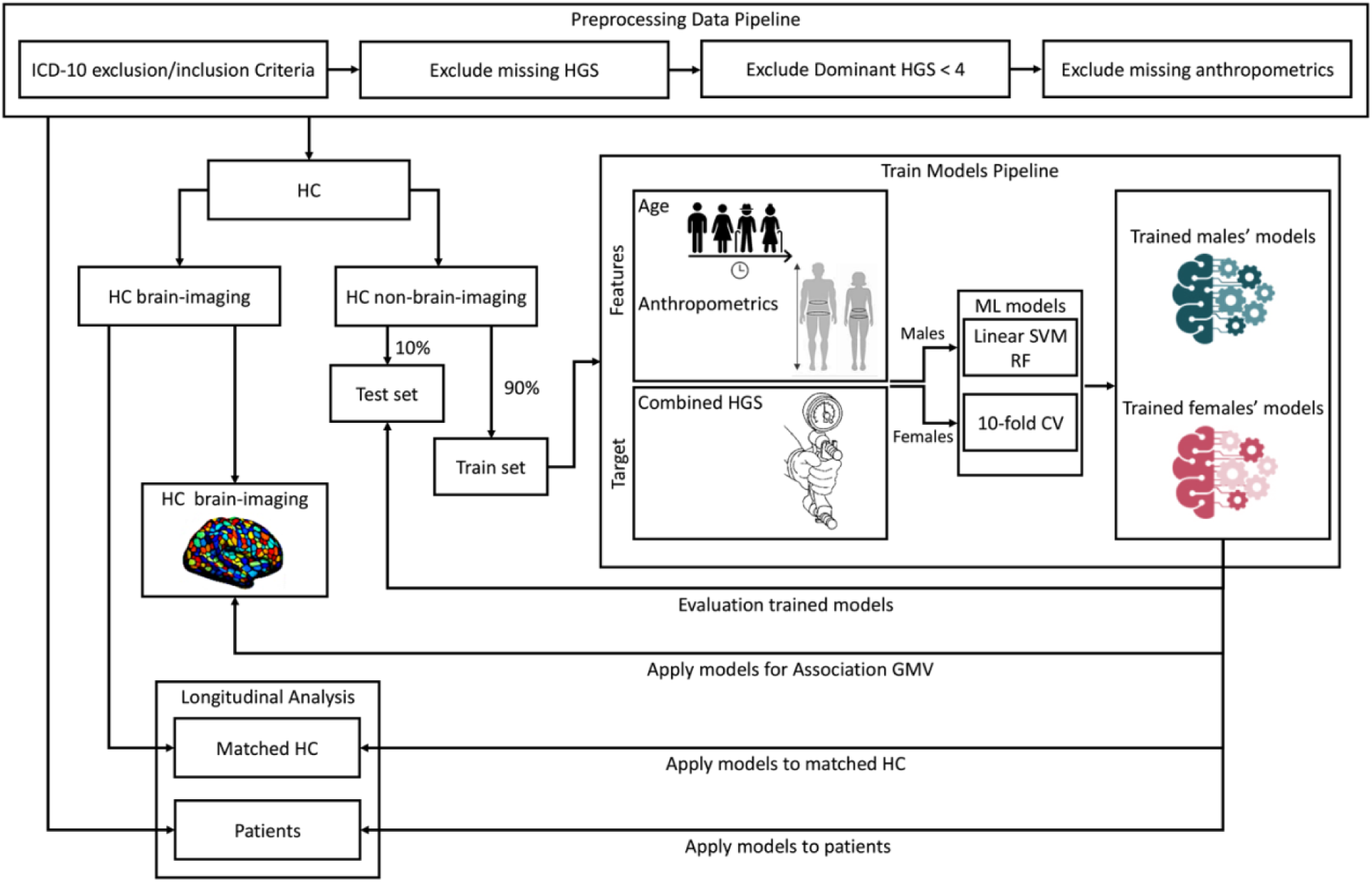
The ML analysis pipeline used to predict HGS and analyze its associations with neurobiological markers and disease-related impairments in this study. For data preparation anthropometric (BMI, height, and waist-to-hip ratio) and demographic (age) variables were obtained from the UK Biobank database. These variables (predictors) were used to train sex-specific ML models, specifically linear SVM and random forest (RF), to predict HGS. The pipeline includes steps for preprocessing, model training, performance evaluation through cross-validation, and the application of statistical bias-correction techniques to enhance prediction accuracy. The true HGS and Δ*HGS* (true HGS – 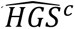; <u>see the “Model training and performance evaluation” section</u>) scores are then assessed for their correlation with neurobiological markers, such as gray matter volume (GMV), and their effectiveness in distinguishing between HC and patients with stroke and MDD.

### Healthy controls

The HC population was obtained by excluding participants with a known history or current diagnosis of mental and behavioral, psychiatric, nervous system, neurological, cardiovascular, cerebrovascular, musculoskeletal system, connective tissue conditions, injuries or poisoning diseases as outlined in the ICD-10 codes (Table 1). The HC participants were divided into two groups: 1) participants who did not undergo brain imaging assessments (non-imaging data, *N* = 201,133) and 2) participants who participated in at least one brain imaging assessment out of the two available imaging assessments (HC with imaging data, *N* = 32,125). Individuals with non-imaging data were used to train and evaluate ML models. The HC individuals with imaging data were employed both in assessing the association between HGS and GMV as well as for matched HC comparisons with patient cohorts (Additional file 1: Figure S1).

**Table 1.**
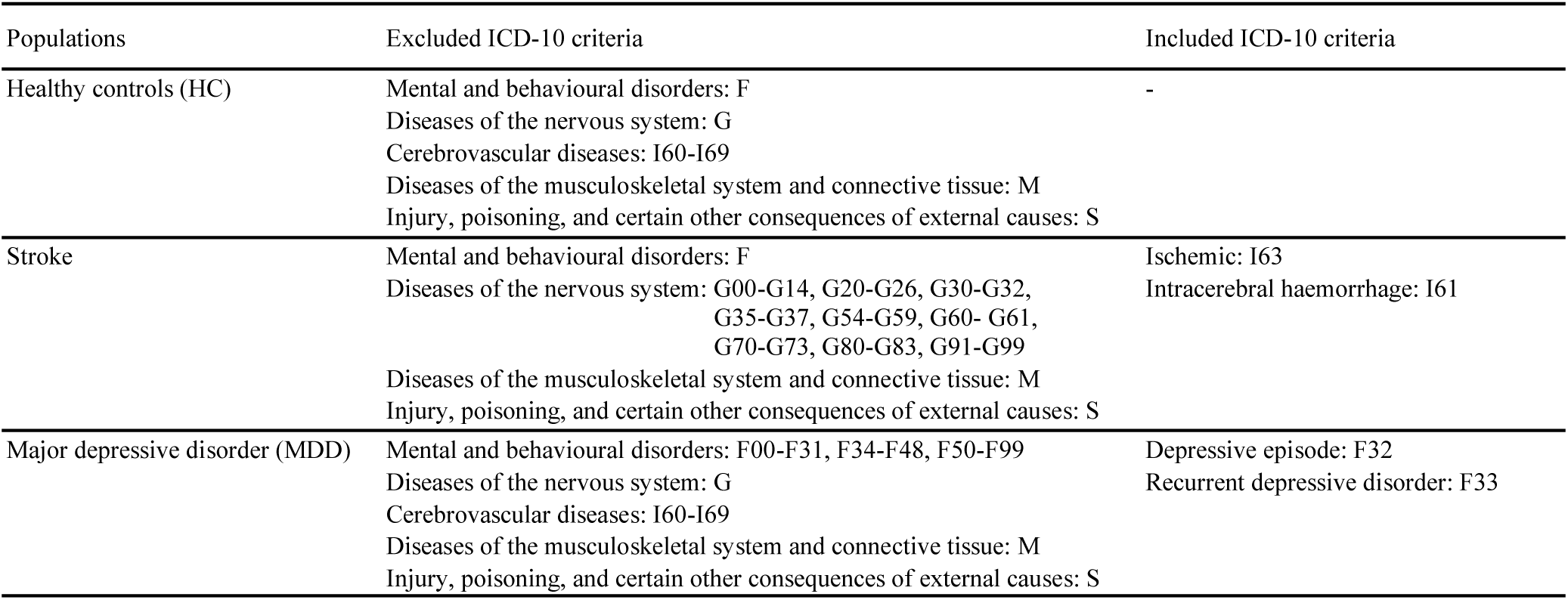
Definition of the study populations based on ICD-10 criteria for HC, stroke, and MDD.

| Populations | Excluded ICD-10 criteria | Included ICD-10 criteria |
| --- | --- | --- |
| Healthy controls (HC) | Mental and behavioural disorders: F<br>Diseases of the nervous system: G<br>Cerebrovascular diseases: I60-I69<br>Diseases of the musculoskeletal system and connective tissue: M<br>Injury, poisoning, and certain other consequences of external causes: S | - |
| Stroke | Mental and behavioural disorders: F<br>Diseases of the nervous system: G00-G14, G20-G26, G30-G32, G35-G37, G54-G59, G60- G61, G70-G73, G80-G83, G91-G99<br>Diseases of the musculoskeletal system and connective tissue: M<br>Injury, poisoning, and certain other consequences of external causes: S | Ischemic: I63<br>Intracerebral haemorrhage: I61 |
| Major depressive disorder (MDD) | Mental and behavioural disorders: F00-F31, F34-F48, F50-F99<br>Diseases of the nervous system: G<br>Cerebrovascular diseases: I60-I69<br>Diseases of the musculoskeletal system and connective tissue: M<br>Injury, poisoning, and certain other consequences of external causes: S | Depressive episode: F32<br>Recurrent depressive disorder: F33 |

### Patient cohorts

First, participants with conditions of the musculoskeletal system, connective tissue, or injury were excluded from all patient sample groups (Table 1). The outcomes of incident stroke were defined according to the “algorithmically-defined outcomes (ADOs)” (UKB Resource 460) developed by the UKB team [29]. The algorithm integrated information from UKB’s baseline assessment data collection along with linkage data, including hospital admissions, diagnoses and procedures, death register records, and self-reported medical condition codes reported at the baseline assessment visit. The incident MDD outcome was obtained from “the first occurrence of health outcomes defined by 3-character ICD-10 code” algorithm (UKB Resource 593) [30,31]. The UK Biobank indicated the first occurrence of a set of diagnostic codes for a wide range of health outcomes across self-report, primary care, hospital inpatient data, and death data, mapped to a 3-digit ICD-10 code. To establish patient cohorts, we included stroke endpoints comprised of ischemic stroke (I63) or intracerebral hemorrhagic stroke (I61), and MDD depressive episode (F32) or recurrent depressive disorder (F33). The onset dates for the diagnoses of two diseases were identified using the first occurrence fields: stroke (Data-Field 42006), and MDD (Data-Fields 130894 and 130896). To ensure diagnostic accuracy, cases based solely on self-reported data were excluded from the analysis. Patients with a history of diseases prior to their baseline assessment visit were excluded to ensure that the analysis focuses on incident cases. Additional exclusion criteria were applied to each patient group, including missing dates of disease onset, missing data, and relevant HGS conditions (see the “Handgrip strength assessment” section). After applying exclusion criteria, we identified disease cohorts consisting of patients with longitudinal data who completed assessments at two time-points: (1) an initial assessment visit prior to the onset of the disease (pre time-point), and (2) the first follow-up assessment visit after disease onset (post time-point). The final patient cohorts comprised: 40 males and 16 females in the stroke group (1 female patient hemorrhagic and all others ischemic), and 37 males and 60 females in the MDD group. These cohorts provided longitudinal data for analysis of HGS changes concerning disease onset.

### Handgrip strength assessment

HGS was measured isometrically using a calibrated Jamar J00105 hydraulic hand dynamometer (Lafayette Instrument Company, USA), which was monitored by a research assistant. During the HGS measurement, participants were told to sit upright in a chair with their forearms resting on armrests pointing forward and their elbows bent and locked at a 90° angle. The maximum HGS value was obtained from each hand while participants were instructed to squeeze the handle as hard as possible for approximately 3 seconds. Both hands were measured consecutively (Data-Field 46 for the left and Data-Field 47 for the right hand). Participants whose dominant HGS < 4 kg or lower than their non-dominant HGS were eliminated from further analysis [1,32]. Hand dominance was based on self-report. If information on handedness was not available or if the individual reported using both hands (ambidextrous) we based dominance on the highest HGS score obtained from either right or left hand. In this study, our target of interest was combined HGS (which we refer to simply as HGS), calculated as the sum of the grip strength measurements from the right and left hands. While measuring HGS in each hand separately can reveal unilateral deficiencies, assessing combined HGS provides a comprehensive measure of overall hand strength. In clinical settings, assessing the strength of both hands offers a robust measure of overall strength and helps identify unilateral weaknesses or conditions affecting one side of the body that may be overlooked with single-hand testing, especially for conditions like stroke or localized musculoskeletal disorders [33]. This approach is particularly relevant when considering the 10% rule, which states that the dominant hand typically has a 10% greater grip strength than the non-dominant hand, primarily applies to right-handed individuals, who make up more than 90% of both male and female participants in this study. In contrast, for left-handed individuals, grip strength tends to be more balanced between both hands [34]. Therefore, combined HGS offers a more equitable assessment for left-handed individuals, as it eliminates the need for adjustments based on hand dominance. Note that for lateralized motor deficits as encountered in stroke, the combined HGS score will also be reduced.

### Demographic and Anthropometric assessments

Participants’ sex was determined from self-reported information (Data-Field 31). Age was calculated based on the date of the baseline assessment attendance and the participant’s birth date. Anthropometric data were obtained during the physical measures phase of each assessment visit. Height (Data-Field 50) was directly measured, while BMI (Data-Field 21001) was calculated using weight and height data (kg/m²). WHR was determined by dividing waist circumference (Data-Field 48) by hip circumference (Data-Field 49).

## ML analysis

### Data preparation

Data from non-imaging HC participants who only attended the baseline assessment was used to train and evaluate ML models. Age and anthropometric (i.e., BMI, height, and WHR) characteristics were considered as predictors in our models. To avoid overfitting and base decisions on the most promising models, we split the HC data into training (90%), and test datasets (10%). The splits were stratified based on binned age (into 5 bins), HGS (into 5 bins), and sex to keep splits representative of the whole population. After excluding cases with missing data and relevant HGS conditions (see the “Handgrip strength assessment” section) from each dataset, the final HC data included 61,816 males (43.32%) and 80,886 females (56.68%) in the training set, and 6,938 males (43.62%) and 8,969 females (56.38%) in the test set (Table 2, Fig. 1, and Additional file 1: Figure S1).

**Table 2.** Summary of the HC non-imaging population characteristics.

| Dataset | HC non-brain-imaging at baseline assessment visit |  |  |  |  |  |
| --- | --- | --- | --- | --- | --- | --- |
|  | Train dataset |  |  | Test dataset |  |  |
| Sex | Both sex | Female | Male | Both sex | Female | Male |
| Number | 142,702 | 80,886 | 61,816 | 15,907 | 8,969 | 6,938 |
| Age, mean (SD) | 55.46 (8.12) | 55.24 (7.99) | 55.75 (8.27) | 55.54 (8.13) | 55.28 (8.01) | 55.87 (8.26) |
| BMI, mean (SD) | 26.76 (4.42) | 26.35 (4.74) | 27.29 (3.91) | 26.74 (4.35) | 26.29 (4.63) | 27.33 (3.89) |
| Height, mean (SD) | 168.46 (9.21) | 162.76 (6.27) | 175.93 (6.79) | 168.47 (9.24) | 162.69 (6.24) | 175.95 (6.8) |
| WHR, mean (SD) | 0.86 (0.09) | 0.81 (0.07) | 0.93 (0.06) | 0.86 (0.09) | 0.81 (0.07) | 0.93 (0.06) |
| Combined HGS, mean (SD) | 62.50 (21.27) | 48.65 (11.68) | 80.62 (16.92) | 62.56 (21.25) | 48.69 (11.69) | 80.48 (16.99) |
| Right dominant hand | 90.34% | 91.93% | 88.26% | 90.16% | 91.6% | 88.3% |

### Model training and performance evaluation

We utilized the non-imaging HC training dataset (61,816 males and 80,886 females) to train sex-specific ML models for HGS prediction using the anthropometrics and age features. To prevent sex bias and given known sex differences in HGS, and anthropometric features, models were trained separately for males and females. We employed linear support vector machine (SVM) and random forest (RF) regression models. Our motivation for including a nonlinear predictive model (RF) besides the linear SVM in our analysis was based on the fact that a non-linear relationship between age and HGS has been already documented [11,20]. Specifically, HGS generally increases until approximately ages 30 to 40, after which it begins to decline, and non-linear models are needed to capture such association [35]. Pearson’s correlation coefficient (*r*), the coefficient of determination (*R*^2^) and mean absolute error (*MAE*) were used to compare model performance. To obtain generalization estimates, we performed 10 times repeated 10-fold (10×10-fold) cross-validation (CV) using the Julearn machine learning library version 0.2.7 (https://juaml.github.io/julearn/) [36], building on top of the scikit-learn library [37]. The hyperparameter *C* for the linear SVM was calculated using a heuristic as 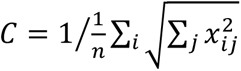 where *n* is the number of subjects [38]. As an alternative, we also trained a RF regression model using the Scikit-Learn (sklearn) Python package version 1.2.1, with 100 trees, a minimum of 2 samples per split, the square root (sqrt) of the total number of features as the maximum number of features considered for the best split, and bootstrapping of the training samples (true) as the hyperparameters (defaults in this version of sklearn).

In the first step, we performed a scaling study by comparing the prediction performances of both the linear SVM and RF models across six levels of sample sizes (10, 20, 40, 60, 80, and 100%) generated by randomly selecting the required number of sampling data from the training dataset to perform the CV procedure. Comparing model performances across different sample sizes help evaluate model stability and reliability by revealing how performance changes with increasing amounts of data. The model with higher performance was selected for further analysis. Feature importance (FI) scores for each model were derived to quantify the influence of individual feature variables on model outputs. For the linear SVM model, we used the coefficient parameter (.coef_) as the FI scores.

In the second step, we validated the models trained using the whole training data (90% of HC), by comparing the true HGS and predicted HGS (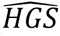) values on the 10% hold-out test set. This step is crucial for determining how well the trained models perform and identifying any inherent biases or errors in the prediction process. To this end, we calculated the difference between the true HGS and the 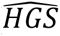 (i.e., 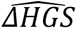 = *true HGS* – 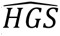). A positive 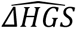 indicates that the individual is stronger than expected, while a negative value suggests weaker than expected strength. However, assessing an individual’s strength using this difference is highly dependent on the accuracy of the HGS prediction model. Prediction frameworks often encounter bias, characterized by overestimation of low values and underestimation of high values in the target variable. Such a bias can compromise model accuracy and impair the interpretability of predictions on new data [39]. To address this issue and enhance model performance, we implemented a statistical bias-correction technique, adjusting HGS predictions to align more closely with the true HGS distribution. We applied the bias-correction method developed by Beheshti et al. for brain age prediction, given its demonstrated effectiveness in reducing variance [40]. The correction is employed by means of a linear regression model between true HGS and the 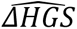. We fitted this model using the predictions from out-of-sample validation sets generated through a 10-fold CV on the training set and calculated the slope (α) and intercept (β) which are used to correct the predictions to achieve a bias-free HGS value of predicted HGS (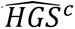).

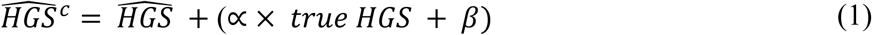

Like the prediction models, the bias correction models were trained separately for females and males. Finally, this 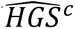 was subtracted from true HGS:

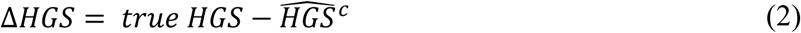

### Reassessment reliability

We then assessed the agreement between Δ*HGS* values by using the reassessment reliability process in the subset of HC test dataset. To evaluate the reliability of the selected sex-specific trained models, we selected participants from the test dataset for which two non-imaging UKB assessment visit sessions were available: (1) baseline assessment visit as the initial measurement, and (2) first repeat assessment visit (after 2-7 years) as the reassessment, resulting in 134 males and 162 females for this analysis (Additional file 1: Table S2). The concordance correlation coefficient (CCC) [41] between Δ*HGS* from the two assessment visit sessions was calculated. This reassessment reliability analysis was designed to assess potential changes in relative HGS over time, considering their new age and possible health changes. The analysis reflects both the stability of scores and their sensitivity to genuine changes in participant conditions over time, which are crucial factors for interpreting the reliability of outcomes.

### Association between brain structure and HGS scores

#### Imaging data and preprocessing

We investigated the neurobiological basis of the HGS scores and their association with GMV. For this, we utilized MRI data from the UKB’s first imaging assessment visit [42], acquired using 3T scanners following the protocol and acquisition parameters detailed in Miller et al. [43]. The structural preprocessing of these images was conducted using pipelines developed and executed by the UKB [44]. Specifically, we analyzed the extracted parcel-wise GMV features from T1-weighted (T1w) preprocessed images. The initial preprocessing of the MRI data involved retrieving T1-weighted preprocessed images from the UK Biobank, which were then converted into a DataLad dataset for provenance tracking [45]. Subsequently, voxel-based morphometry was computed using the Computational Anatomy Toolbox (CAT) version 12.7 resulting in images normalized to the MNI152 space with a 1.5 mm isotropic resolution [46]. The parcel-wise GMV was extracted as the winsorized mean (with limits set at 10%) of the voxel-wise values per parcel, combining three different brain atlases: the Schaefer et al. cortical atlas (1000-parcel) [47], the S4 3T version of Tian et al. Melbourne subcortical atlas (54-parcel) [48], and the cerebellum SUIT Diedrichsen et al. atlas (34-parcel) [49]. The result was a feature vector containing the parcellated GMV of 1,088 brain regions of interest (ROI) for each participant. To accommodate for individual differences in total intracranial volume (TIV), we linearly regressed it out from each brain region. For this analysis, we used a subset of HC participants who completed the initial imaging visit with 11,077 males and 12,849 females (Additional file 1: Table S1). The final samples were reduced to 7,726 males and 9,292 females based on the availability of all 1,088 GMV features.

#### Correlation analysis

We investigated the correlation between the GMV of 1,088 regions separately for true HGS and Δ*HGS*. The Δ*HGS* values were obtained using the best-performing separate linear SVM models for males and females (see the “Model training and performance evaluation” section). Pearson’s *r* and corresponding p-values were computed. To focus on robust associations, we applied a correlation threshold of |*r*| > 0.1 (the absolute values of correlation coefficients exceeding 0.1) together with correcting *p*-values using the Bonferroni correction with significance determined at *p*_*corrected*_ < 0.05. Separate analyses were conducted for female and male participants to identify regions significantly associated with each HGS score. The correlation coefficients for these significant regions were visualized on brain maps for each sex individually. Additionally, the intersection of the regions showing significance for both sexes were visualized using averaged correlation coefficients.

### Evaluation of pre-to-post disease longitudinal HGS changes

#### Data preparation

We identified patient cohorts with longitudinal data from pre (prior to the disease onset) and post (follow-up the disease onset) time-points. The Δ*HGS* score could provide insights into disease-specific variation in HGS between pre and post time-points on longitudinal cases. To investigate this, patients with diseases were compared with matched HC samples using a 1:10 (case:control) ratio, ensuring robust comparative analyses. The matching process entailed selecting HC individuals whose assessment visit sessions at both pre and post time points coincided with those of the patients, thereby maintaining temporal consistency.

The final patient cohorts consisted of 40 males and 16 females with stroke, and 37 males and 60 females with MDD (see the “Patient cohorts” section). Matched HCs were selected from the pool of HC participants who had undergone brain imaging assessment visits and were not used in model training (15,516 males and 16,609 females). Further refinement of subsamples involved excluding data with missing values and additional HGS conditions (see the “Handgrip strength assessment” section). The final number of HC participants included in the matching process for each assessment was as follows: baseline (11,918 male and 13,714 female), first repeat (1,884 male and 2,013 female), imaging visit (11,077 male and 12,849 female), and first repeat imaging (1,153 males and 1,359 females). These refined HC cohorts enabled comprehensive comparisons with patient cohorts across multiple assessment time points. The HC subsample used in the matched control-case study, categorized by assessment visit, is detailed in Additional file 1: Table S1. Propensity score matching (PSM) was applied using a 1:10 nearest-neighbor approach to select HC samples for each patient within each disease cohort [50]. This method involved matching each patient with 10 HC participants who had similar propensity scores, taking into account age, anthropometric features at the pre time-point, and the time interval between pre- and post-assessment visits (days). Finally, post time-point data were identified for each HC participant to maintain subject consistency between pre and post time-points. This approach ensured the availability of longitudinal data in the matched HC group, enabling a robust temporal comparison with the patient cohorts. The matching process was conducted without replacement, ensuring that each HC individual could be selected as a match only once per patient group (an overlap was allowed of HC for the different patient samples, i.e., overlaps of HC for stroke and HC for MDD). The characteristics of the matched HC samples and patients summarized in Table 3.

**Table 3.** Characteristics of matched healthy controls (HC) and patients.

| Sex | Male |  |  |  | Female |  |  |  |
| --- | --- | --- | --- | --- | --- | --- | --- | --- |
|  | Patient |  | Matched HC |  | Patient |  | Matched HC |  |
|  | Pre | Post | Pre | Post | Pre | Post | Pre | Post |
| **Stroke** |  |  |  |  |  |  |  |  |
| Number | 40 | 40 | 400 | 400 | 16 | 16 | 160 | 160 |
| Age, mean (SD) | 59.65 (6.79) | 68.91 (7.95) | 59.31 (6.86) | 68.24 (7.34) | 61.72 (5.08) | 71.21 (6.91) | 61.29 (5.84) | 71.01 (7.34) |
| BMI, mean (SD) | 26.7 (3.23) | 25.85 (3.32) | 26.68 (3.79) | 26.53 (4.02) | 26.37 (5.26) | 26.7 (5.36) | 26.68 (4.71) | 26.36 (4.79) |
| Height, mean (SD) | 177.22 (7.23) | 176.44 (7.19) | 177.34 (6.35) | 176.71 (6.51) | 162.39 (7.43) | 161.3 (7.02) | 162.32 (5.89) | 161.34 (6.05) |
| WHR, mean (SD) | 0.94 (0.06) | 0.94 (0.06) | 0.94 (0.06) | 0.94 (0.06) | 0.82 (0.1) | 0.84 (0.1) | 0.82 (0.07) | 0.84 (0.07) |
| Combined HGS, mean (SD) | 77.55 (16.45) | 70.75 (12.9) | 80.79 (15.83) | 68.84 (16.57) | 44.31 (13.03) | 44.12 (12.85) | 47.08 (11.08) | 42.34 (11.4) |
| Right dominant hand (%) | 37 (92.5%) | 37 (92.5 %) | 359 (89.75%) | 359 (89.75%) | 16 (100%) | 16 (100%) | 154 (96.25 %) | 154 (96.25 %) |
| Time to/from disease onset, mean (SD) | -5.14 (2.87) | 4.12 (3.28) |  |  | -5.71 (3.57) | 3.79 (3.13) |  |  |
| Time elapsed pre- to post- (years) |  | 9.26 |  | 8.93 |  | 9.5 |  | 9.72 |
| **MDD** |  |  |  |  |  |  |  |  |
| Number | 37 | 37 | 370 | 370 | 60 | 60 | 600 | 600 |
| Age, mean (SD) | 54.9 (6.89) | 64.36 (6.82) | 55.76 (7.84) | 65 (7.77) | 52.72 (7.55) | 62.02 (7.91) | 52.8 (7.53) | 62.02 (7.9) |
| BMI, mean (SD) | 27.38 (4.14) | 28.21 (4.55) | 27.58 (4.05) | 27.37 (4.18) | 26.01 (3.83) | 26.8 (5.03) | 26.04 (4.8) | 26 (5.07) |
| Height, mean (SD) | 176.02 (6.14) | 175.49 (6.16) | 176.13 (6.44) | 175.7 (6.57) | 163.01 (5.09) | 162.45 (4.99) | 163.28 (6.27) | 162.8 (6.3) |
| WHR, mean (SD) | 0.93 (0.05) | 0.97 (0.05) | 0.93 (0.06) | 0.94 (0.07) | 0.79 (0.06) | 0.82 (0.07) | 0.79 (0.06) | 0.81 (0.07) |
| Combined HGS, mean (SD) | 78.89 (14.48) | 72.59 (14.06) | 82.44 (16.04) | 70.77 (17.06) | 49.98 (12.01) | 45.48 (12.2) | 53.16 (11.41) | 43.73 (11.91) |
| Right dominant hand (%) | 34 (91.89%) | 34 (91.89%) | 328 (88.65%) | 328 (88.65%) | 56 (93.33%) | 56 (93.33%) | 573 (95.5%) | 573 (95.5%) |
| Time to/from disease onset, mean (SD) | -5.78 (3.35) | 3.69 (2.6) |  |  | -5.26 (3.38) | 4.04 (3.13) |  |  |
| Time elapsed pre- to post- (years) |  | 9.46 |  | 9.24 |  | 9.3 |  | 9.22 |

### Statistical analysis

The study employed ANOVAs (Analysis of Variance) to compare each of the HGS scores, true HGS and Δ*HGS*, between each patient group (i.e., stroke and MDD) and their corresponding matched HC samples. The Δ*HGS* values were obtained using the best-performing linear SVM models separate for males and females, detailed in the “Model training and performance evaluation” section. The assumptions of the ANOVAs were evaluated using Kolmogorov-Smirnov (KS) normality test and Levene’s test (testing variance homogeneity). The assumption of normality was not met for all patient groups. However, extensive literature supports that ANOVA is generally robust to violations of this assumption, particularly with larger sample sizes. For samples larger than 12, non-normality has minimal impact on Type I error rates. Furthermore, with samples >50 per group, the central limit theorem enhances the robustness of the analysis [51–54]. Consequently, the violation of normality in our analysis is unlikely to significantly affect the validity of the ANOVA results. While homogeneity of variance was met for the factor diagnosis for all patient groups, for the factor sex the assumption was mostly violated. Consequently, two-way ANOVA was conducted separately for each sex, with two main factors: time-point (pre- and post-) as a “within-subject” factor for repeated scores, and the health condition considered as a “between-subject” factor. For significant interactions, Tukey-Kramer post-hoc analysis was used to compare groups separately for each time-point and within each group.

## Results

### HGS predictive modelling

We found that HGS can be predicted using both machine learning models, i.e., linear SVM and RF. However, the linear SVM outperformed RF across all sample sizes, achieving higher performance metrics measured as Pearson’s correlation coefficient (*r*), *R*^2^, and MAE for both male and female cohorts (Fig. 2, Table 4). Both models showed a decrease in variance with increasing sample size, as expected. The average performance stabilized after using 40% of the data indicating that the sample size used is large enough to capture the predictive signal adequately.

**Fig. 2.**
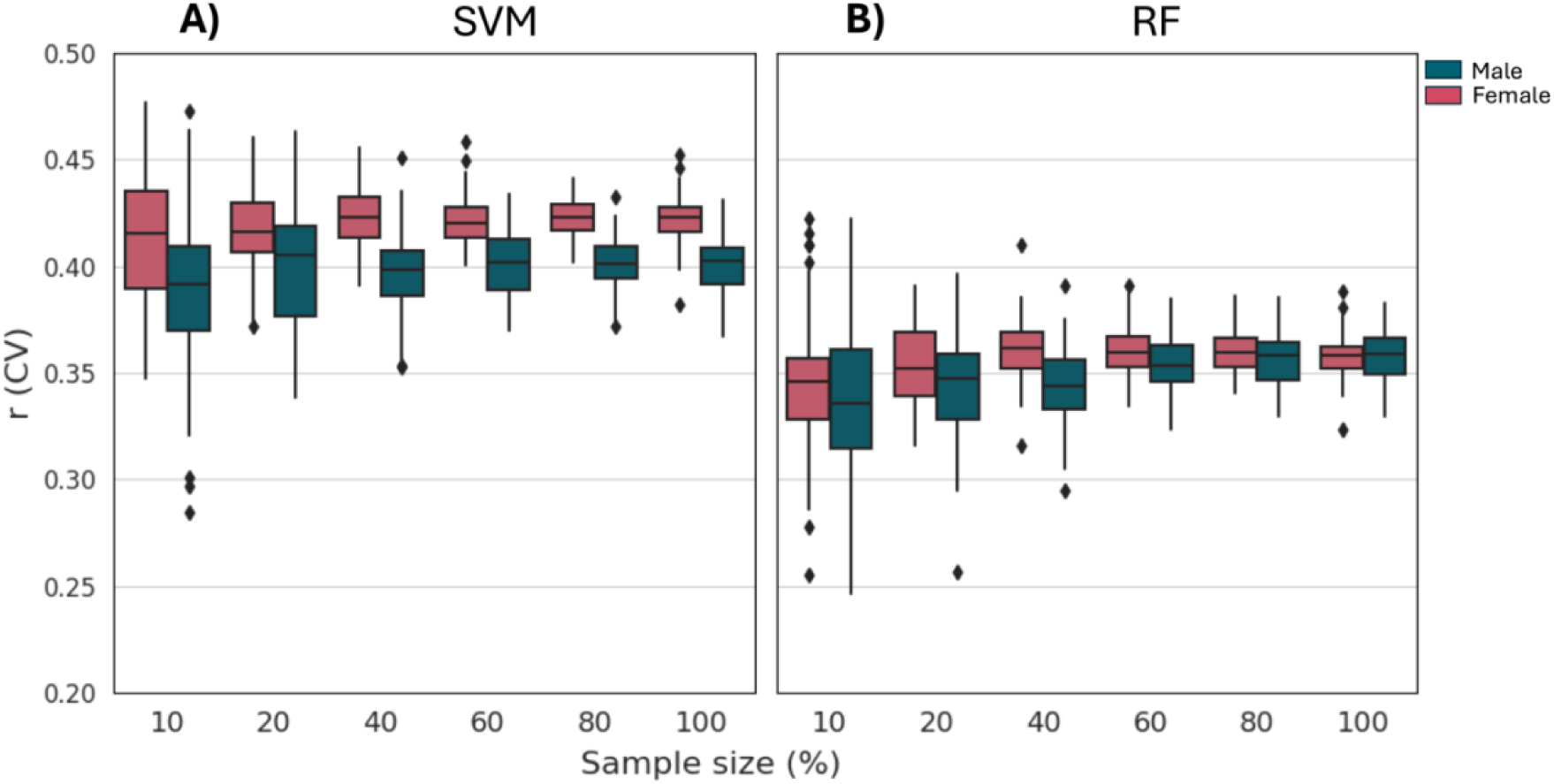
Pearson correlation coefficient (r) from CV analysis at different sample sizes. **A)** linear SVM and **B)** RF models built on the training dataset with increasing sample sizes (10% to 100 %). The sample size indicates the percentage of data used for performing CV. In males, SVM achieved a maximum median *r* of 0.405 and *R*^2^ of 0.163 compared to the RF’s 0.347 and 0.096 at 20% of the sample size (*N* = 12,364). In females, SVM had a maximum median *r* of 0.423 and *R*^2^ of 0.179 compared to the RF’s 0.359 and 0.107 at 80% of the sample size (*N* = 64,712).

**Table 4.**
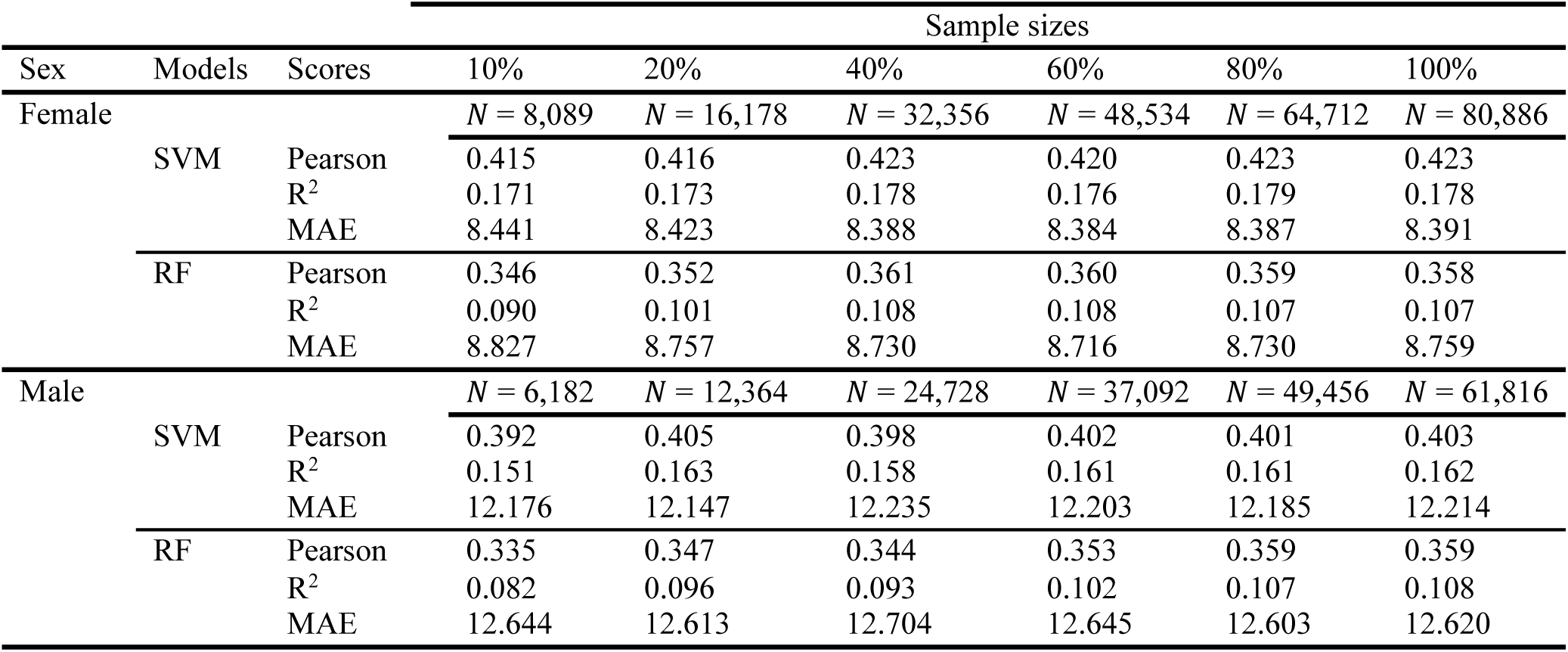
Comparison of linear SVM and RF models by increasing sample size on the training dataset.

### Model Validation

To validate the selected linear SVM models, which were separately trained on the whole training dataset for males and females. Then we applied them to the independent 10% HC test dataset, comprising 6,938 males and 8,969 females. After applying bias-correction to the predictions (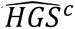) on the test dataset, prediction accuracy significantly improved compared to the uncorrected predictions (Additional file 1: Figure S2). For males, the Pearson’s correlation coefficients increased from *r* = 0.40 (without bias-correction) to *r* = 0.94 (with bias-correction, *p* < 0.0001), and for females, it improved from *r* = 0.42 (without bias-correction) to *r* = 0.93 (with bias-correction, *p* < 0.0001), demonstrating enhanced performance (Fig. 3A). The bias-correction was performed using the method proposed by Beheshti et al. [40], involving the calculation of slope (α) and intercept (β) by fitting a linear regression model between true HGS and the residuals (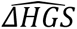 = *true HGS* – 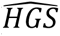). Bias-correction parameters were estimated separately for males (α = 0.839, β = −67.631) and females (α = 0.822, β = −39.971) using predictions from out-of-sample validation sets generated via 10-fold CV on the HC training dataset. We then assessed the models’ unbiasedness by examining the correlation between *ΔHGS* (*true* HGS – 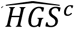) and true HGS (Fig. 3B). The analysis revealed no significant correlation between Δ*HGS* and true HGS for either males (*r* = 0.01) or females (*r* = 0.00), indicating an absence of bias in the corrected predictions. Before bias-correction, correlations were *r* = 0.92 for males and *r* = 0.91 for females, indicating some initial bias. This finding supports the conclusion that the model is unbiased across both sexes.

**Fig. 3.**
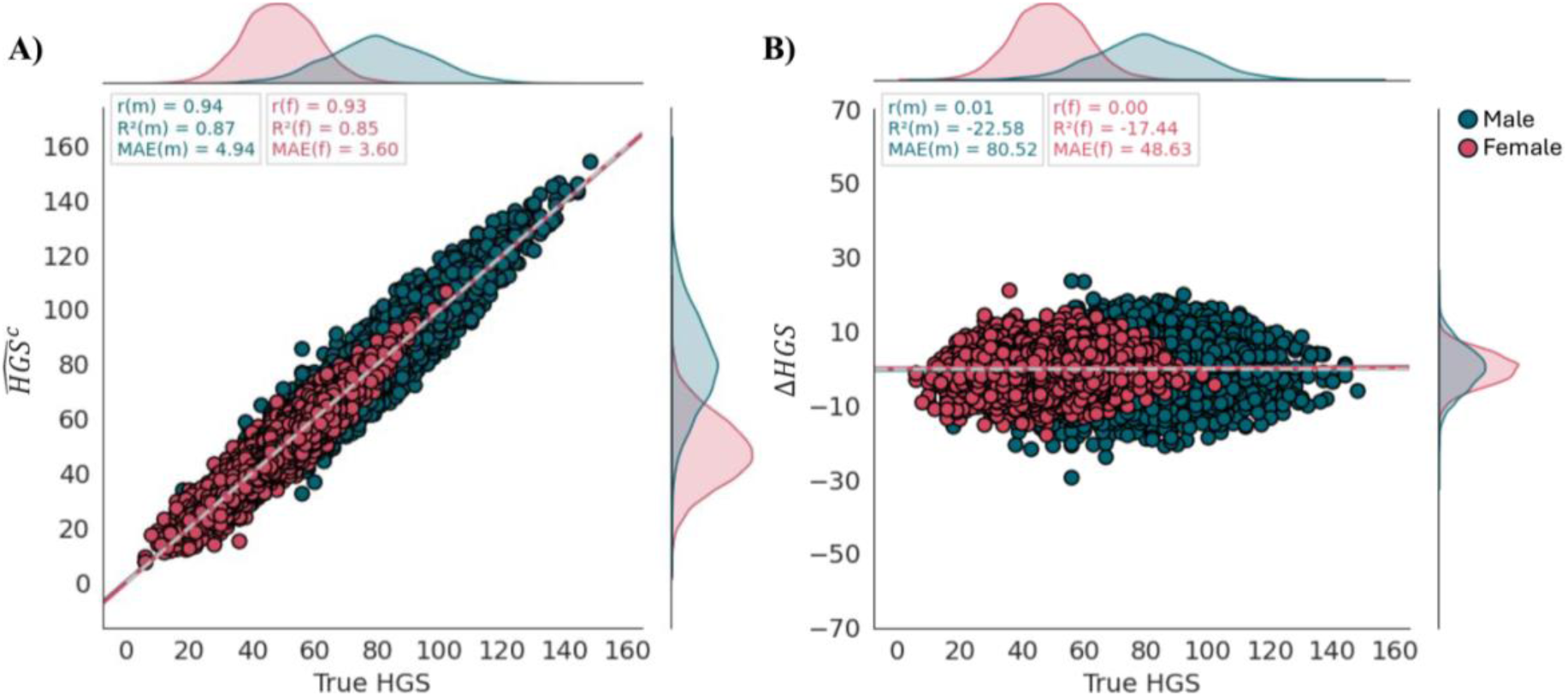
Relationship between 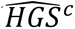 (A) and ΔHGS (B) scores versus true HGS, respectively, after applying bias-correction method on the independent non-brain-imaging HC test dataset, for males (N = 6,938) and females (N = 8,969). **A)** Scatter plot of 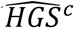 and true HGS: for males (r = 0.94, R^2^ = 0.87, MAE = 4.94) and for females (r = 0.93, R^2^ = 0.85, MAE = 3.6). **B)** Scatter plot of ΔHGS and true HGS: for males (r = 0.01, p = 0.41) and for females (r = 0.00, p = 0.73). The dashed grey line in the A indicates the identity line (y = x), while the dashed grey line in the B indicates the reference line (y = 0).

We then investigated the FI scores from the final models trained on the training dataset (90% of HC). The models showed similarities and differences between males and females. Although the direction of the contribution was the same for both sexes, the strength of contributing factors differed. For males, the linear SVM model identified height and BMI as having positive contributions to HGS (FI_height_ = 4.94 and FI_BMI_ = 3.00), while WHR (FI_WHR_ = -2.57) and age (FI_age_ = -2.83) showed negative coefficients. In females, height (FI_height_ = 3.27) and BMI (FI_BMI_ = 0.67) remained positive contributors, and WHR (FI_WHR_ = -0.53) and age (FI_age_ = -3.07) showed negative effects.

### Reassessment reliability of ΔHGS

The reassessment reliability was evaluated using the concordance correlation coefficient (CCC) between Δ*HGS* values for two sessions: the baseline assessment visit as the initial session (session 0), and the first repeat assessment visit (after 2-7 years) as reassessment (session 1) on the same participants from the non-brain-imaging HC test dataset (*N* = 296, 54.73% female) without and with bias-correction. The results demonstrated high reassessment reliability for Δ*HGS* after applying bias-correction, with males showing a CCC of 0.90 and females a CCC of 0.89. These values were notably higher compared to the Δ*HGS* without bias-correction (Table 5). The high reassessment reliability indicates that Δ*HGS* scores remain stable despite physiological changes in participants’ conditions over 2-7 years. This reliability is crucial for application in the elderly population, where age and health-related changes considerably affect HGS. The strong agreement between sessions, shown by high CCC values, confirms the reliability of the sex-specific models.

**Table 5.** CCC between baseline and follow-up delta values, and MAE with and without bias correction.

| Sex | N | Without bias-correction |  |  | With bias-correction |  |  |
| --- | --- | --- | --- | --- | --- | --- | --- |
|  |  | MAE (Session 0) | MAE (Session 1) | CCC | MAE (Session 0) | MAE (Session 1) | CCC |
| Female | 162 | 8.129 | 12.059 | 0.32 | 3.294 | 3.496 | 0.89 |
| Male | 134 | 12.934 | 16.434 | 0.47 | 4.534 | 4.475 | 0.90 |

### Association between HGS scores and brain structure

We investigated Pearson’s *r* between regional GMV and two HGS scores, true HGS and Δ*HGS* in a large cohort of HC (7,726 males and 9,292 females). Each participant’s data included 1,088 regional GMV features covering the whole brain (see the “Imaging data and preprocessing” section). Significant correlations were identified for both true HGS and Δ*HGS* after applying Bonferroni correction (*p*_corrected_ < 0.05). Fig. 4 demonstrates the distribution of associations between regional GMV and HGS. The cortical region labeling was derived from the Schaefer 1000-parcel 7-network brain atlas.

**Fig. 4.**
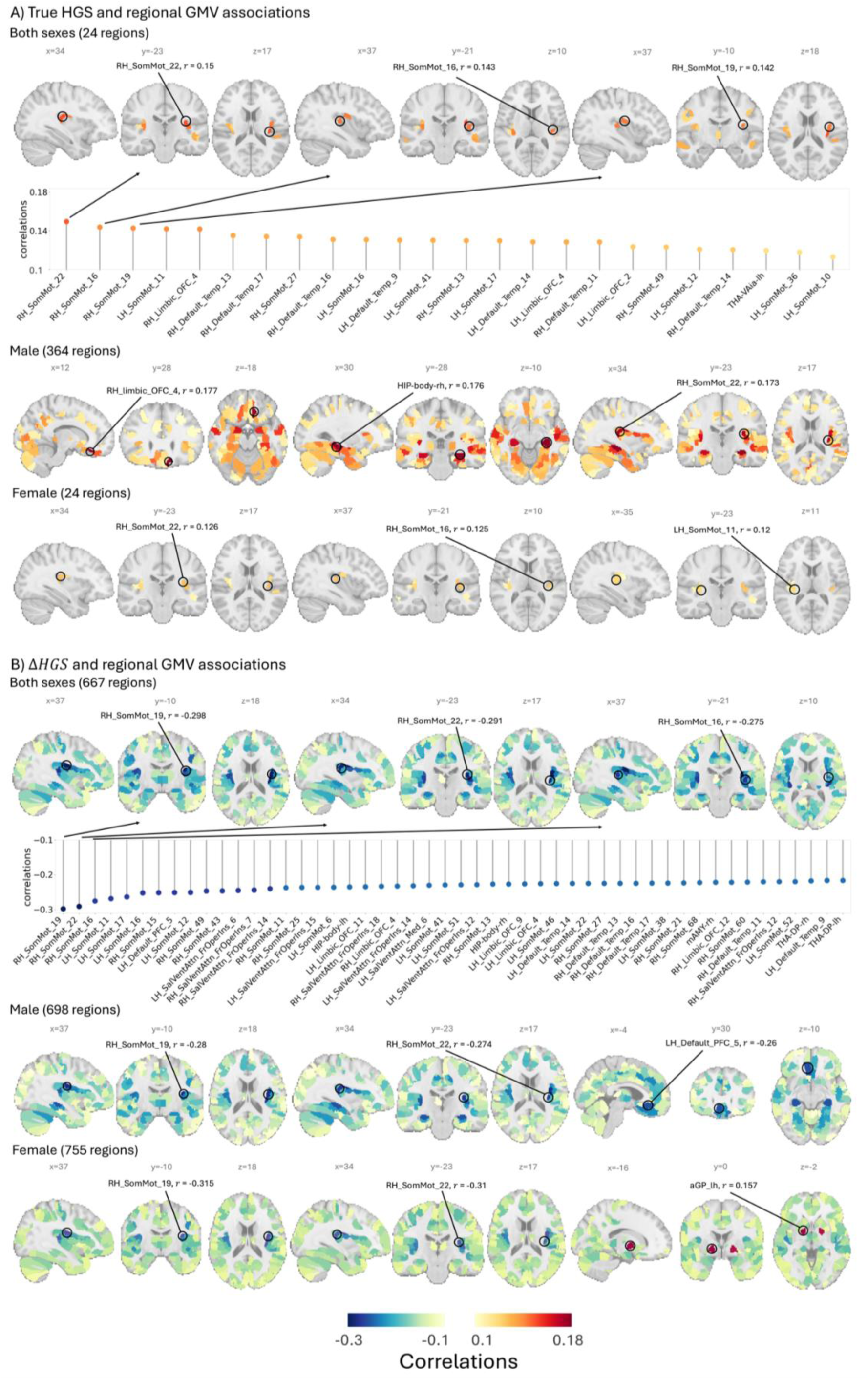
Regional distribution of associations between GMV and HGS, after applying Bonferroni correction (*p*_*corrected* < 0.05) and focusing on significant regions with |*r*| > 0.1. **A) True HGS and GMV correlation:** Both sexes (24 regions): The average correlation between intersecting of significant regions in both sexes ranged (0.113≤ *r* ≤0.15). The strongest cortical correlation (*r* = 0.15) was observed in “RH_SomMot_22”, the strongest subcortical correlation (*r* = 0.12) in “THA-VAia-lh”. No cerebellar regions were identified among these 24 regions. The scatter plot represents the correlation of these 24 significant regions. Males (364 regions): Correlations ranged (0.1≤ *r* ≤0.177), with the strongest cortical correlation (*r* = 0.177) in “RH_Limbic_OFC_4”, the strongest subcortical correlation (*r* = 0.176) in “HIP-body-rh”, and the highest cerebellar correlation (*r* = 0.133) in “Left_VI”. Females (24 regions): Correlations ranged (0.1≤ *r* ≤0.126), with the strongest cortical correlation (*r* = 0.126) in “RH_SomMot_22”, the strongest subcortical correlation (*r* = 0.113) in “THA-VAia-lh”, and no cerebellar regions were significant. **B) ΔHGS and GMV correlation:** Both sexes (667 regions): The average correlations between the intersecting of significant regions in both sexes ranged (-0.298≤ *r* ≤-0.101), with the strongest cortical correlation (*r* = -0.298) in “RH_SomMot_19”, the strongest subcortical correlation (*r* = -0.235) in “HIP-body-lh”, and the highest cerebellar correlation (*r* = -0.214) in “Left_VI”. The scatter plot represents the 50 top significant regions. Males (698 regions): Correlations ranged (-0.28≤ *r* ≤-0.1), with the strongest cortical correlation (*r* = -0.28) in “RH_SomMot_19”, the highest subcortical association (*r* = -0.256) in the “HIP-body-rh”, and the highest cerebellar correlation (*r* = -0.222) in the “Left_VI”. Females (755 regions): Correlations ranged (-0.315≤ *r* ≤0.157), with the strongest cortical correlation (*r* = -0.315) in “RH_somMot_19”, the strongest subcortical correlation (*r* = -0.231) in “THA-DP-rh”, the highest cerebellar correlation (*r* = -0.205) in “Left_VI”. Two subcortical regions, “aGP_lh” (*r* = 0.157) and “aGP_rh” (*r* = 0.145), showed positive correlations.

For true HGS, significant correlations were found in 878 regions for males and 660 for females. All correlations were positive. When applying a correlation threshold of |*r*| > 0.1, 364 regions remained in males and 24 in females. The intersection of significant regions between sexes included 24 regions, none of which were located in cerebellar areas. These results indicate a broader association between true HGS and GMV in males compared to females, suggesting potential sex differences. The average correlations between true HGS and regional GMV of the intersecting significant regions for both sexes showed values ranging from 0.113 to 0.15 (Fig. 4A). The strongest correlations were observed in the cortical right hemisphere somatomotor regions, parcels 22 (RH_SomMot_22, *r* = 0.15), 16 (RH_SomMot_16, *r* = 0.143), and 19 (RH_SomMot_19, *r* = 0.142). Subcortical associations differed across sexes (Additional file 1: Table S3), with the highest subcortical association found in the left hemisphere inferior ventral anterior division of the thalamus (THA-VAia-lh, *r* = 0.12). The results showed that a higher HGS is linked with increased GMV in the cortical areas, which are crucial for motor control.

In contrast, Δ*HGS* demonstrated significant correlations in 968 regions for males and 993 regions for females. After applying the threshold of |*r*| > 0.1, 698 regions in males and 755 regions in females remained. The intersection of significant regions between sexes retained 667 regions, highlighting the consistency and robustness of the association between the Δ*HGS* score and GMV. The average correlations for these intersecting significant regions ranged from -0.298 to -0.101 (Fig. 4B). The only positive correlations were observed in females within subcortical regions, specifically in the anterior globus pallidus of the left hemisphere (aGP_lh, *r* = 0.157) and the right hemisphere (aGP_rh, *r* = 0.145). For both sexes, the strong associations included the right hemisphere somatomotor cortical region from the somatosensory network, parcel 19 (RH_SomMot_19, *r* = -0.298), followed by parcel 22 (RH_SomMot_22, *r* = -0.291), and parcel 16 (RH_SomMot_16, *r* = -0.275). This pattern persisted in sex-specific analyses, with males showing strong correlations in RH_SomMot_19 (*r* = -0.28) and RH_SomMot_22 (*r* = -0.274), while females demonstrated even stronger correlations in these regions (RH_SomMot_19: *r* = -0.315; RH_SomMot_22: *r* = -0.307). Subcortical associations differed across sexes (Additional file 1: Table S4), with the highest subcortical association in both sexes was found in the left hemisphere hippocampal body (HIP-body-lh, *r* = -0.235). The most significant cerebellar association was observed in the left lobule VI (Left_VI) for both sexes (*r* = -0.214). The higher number of significant and stronger correlations observed across a broader range of brain areas suggest that Δ*HGS* can be a more sensitive score for detecting brain structure relationships. Furthermore, the significant increase in overlapping regions suggested a common neurobiological basis beyond sex differences.

### Evaluation of pre-to-post disease longitudinal HGS changes

The innovative aspect of this study is the implementation of a longitudinal design, which captures HGS scores at two critical time points: before (pre) and after (post) disease onset. Capturing longitudinal changes in HGS can improve our understanding of the dynamic relationship between HGS and disease onset and progression, providing valuable insights into how disease impacts physical function over time. Such changes can provide insights into an individual’s health status, the effectiveness of training or rehabilitation programs, and the progression of various health conditions. To analyze this longitudinal data, we employed a two-way ANOVA, separately for each sex. The analyses included group (patient/control) as the between-subject factor and time-point (pre/post) as the within-subject factor, with either true HGS or Δ*HGS* as the dependent scores. We analyzed the stroke and MDD cohorts separately with 10 matched HC for each patient (Table 3).

A significant main effect of time-point was observed for both patient cohorts, indicating time-dependent changes in HGS and Δ*HGS* across patients and healthy controls (Table 6). In stroke patients, a significant interaction was observed in males for both true HGS (*F*_1,438_ = 4.434, *p* = 0.036) and Δ*HGS* (*F*_1,438_ = 9.91, *p* = 0.002). Post-hoc analysis revealed the significant differences in matched HC between pre and post time-points for both scores (*p* < 0.0000), and in patients between pre and post time-points for Δ*HGS* score (*p* = 0.0332). No significant group differences were found at any time point, and no significant interactions were observed in female stroke patients (Fig. 5).

**Fig. 5.**
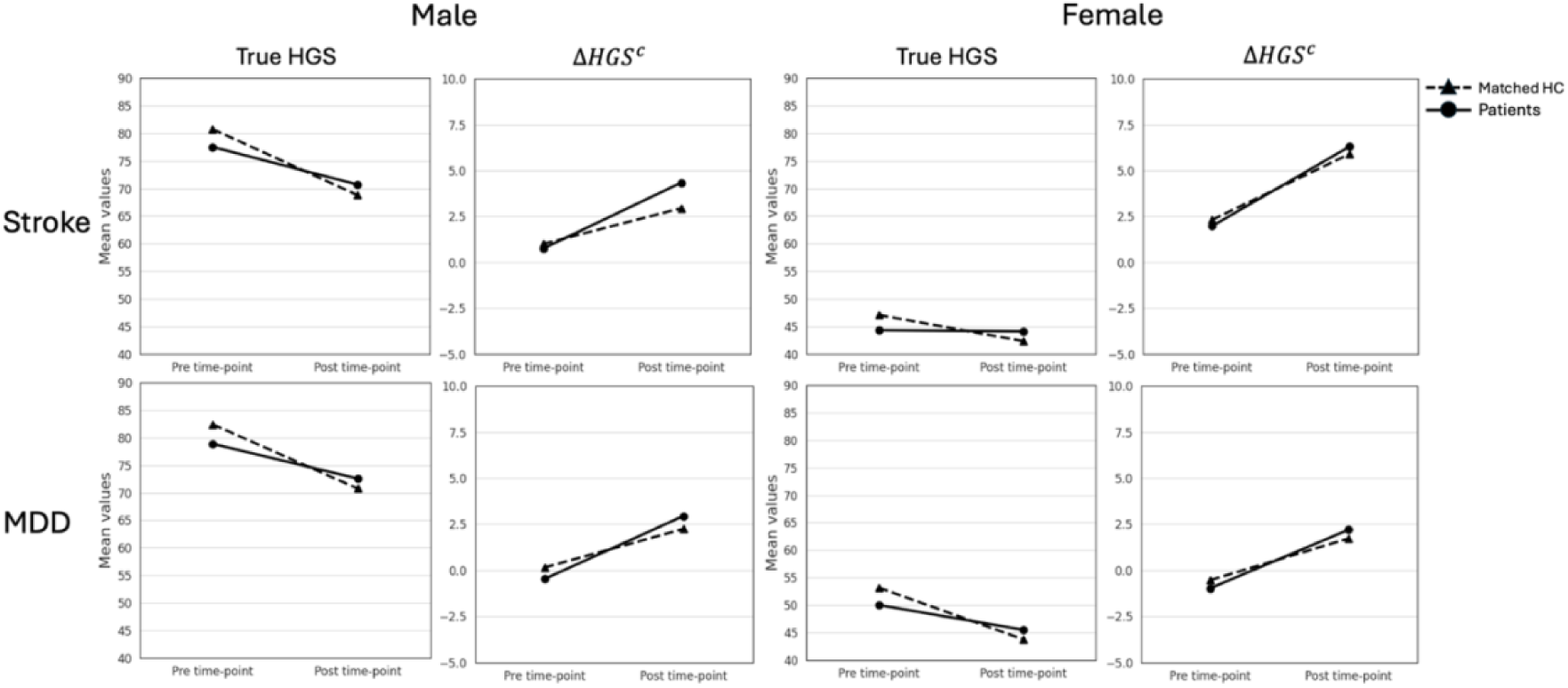
Changes in HGS among patients with stroke and MDD compared to matched HC across two time-points: pre and post time-points. The dashed lines represent the matched control group, while the solid lines depict the patient groups. These findings underscore the importance of considering both absolute and relative scores of muscle strength in understanding the physical capabilities of individuals with neurological and psychiatric diseases. The increased Δ*HGS* suggests that despite the decline in true HGS, patients and matched HC may perform better than expected when accounting for baseline physical attributes.

**Table 6.**
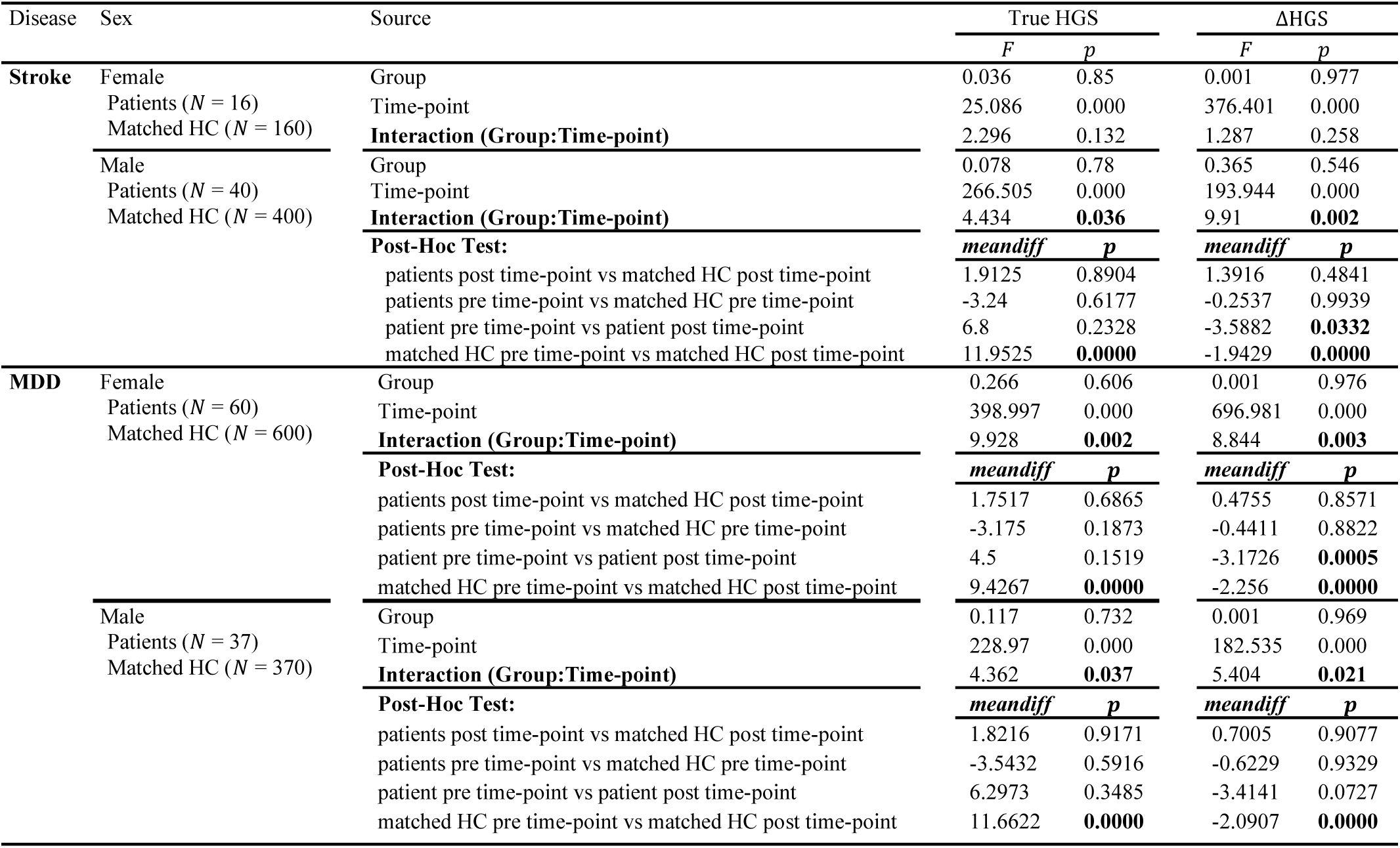
Sex-specific ANOVA and post-hoc results for Group (matched HC vs. patient) and Time-point (pre, post)

| Disease | Sex | Source | True HGS | | $\Delta HGS$ | |
| --- | --- | --- | --- | --- | --- | --- |
|  |  |  | *F* | *p* | *F* | *p* |
| Stroke | Female | Group | 0.036 | 0.85 | 0.001 | 0.977 |
|  |  | Time-point | 25.086 | 0.000 | 376.401 | 0.000 |
|  |  | Interaction (Group:Time-point) | 2.296 | 0.132 | 1.287 | 0.258 |
|  | Male | Group | 0.078 | 0.78 | 0.365 | 0.546 |
|  |  | Time-point | 266.505 | 0.000 | 193.944 | 0.000 |
|  |  | Interaction (Group:Time-point) | 4.434 | **0.036** | 9.91 | **0.002** |
|  |  | Post-Hoc Test: | *meandiff* | *p* | *meandiff* | *p* |
|  |  | patients post time-point vs matched HC post time-point | 1.9125 | 0.8904 | 1.3916 | 0.4841 |
|  |  | patients pre time-point vs matched HC pre time-point | -3.24 | 0.6177 | -0.2537 | 0.9939 |
|  |  | patient pre time-point vs patient post time-point | 6.8 | 0.2328 | -3.5882 | **0.0332** |
|  |  | matched HC pre time-point vs matched HC post time-point | 11.9525 | **0.0000** | -1.9429 | **0.0000** |
| MDD | Female | Group | 0.266 | 0.606 | 0.001 | 0.976 |
|  |  | Time-point | 398.997 | 0.000 | 696.981 | 0.000 |
|  |  | Interaction (Group:Time-point) | 9.928 | **0.002** | 8.844 | **0.003** |
|  |  | Post-Hoc Test: | *meandiff* | *p* | *meandiff* | *p* |
|  |  | patients post time-point vs matched HC post time-point | 1.7517 | 0.6865 | 0.4755 | 0.8571 |
|  |  | patients pre time-point vs matched HC pre time-point | -3.175 | 0.1873 | -0.4411 | 0.8822 |
|  |  | patient pre time-point vs patient post time-point | 4.5 | 0.1519 | -3.1726 | **0.0005** |
|  |  | matched HC pre time-point vs matched HC post time-point | 9.4267 | **0.0000** | -2.256 | **0.0000** |
|  | Male | Group | 0.117 | 0.732 | 0.001 | 0.969 |
|  |  | Time-point | 228.97 | 0.000 | 182.535 | 0.000 |
|  |  | Interaction (Group:Time-point) | 4.362 | **0.037** | 5.404 | **0.021** |
|  |  | Post-Hoc Test: | *meandiff* | *p* | *meandiff* | *p* |
|  |  | patients post time-point vs matched HC post time-point | 1.8216 | 0.9171 | 0.7005 | 0.9077 |
|  |  | patients pre time-point vs matched HC pre time-point | -3.5432 | 0.5916 | -0.6229 | 0.9329 |
|  |  | patient pre time-point vs patient post time-point | 6.2973 | 0.3485 | -3.4141 | 0.0727 |
|  |  | matched HC pre time-point vs matched HC post time-point | 11.6622 | **0.0000** | -2.0907 | **0.0000** |

In MDD patients, significant interactions were observed in both females and males for Δ*HGS* (males: *F*_1,405_ = 5.404, *p* = 0.021; females: *F*_1,658_ = 8.844, *p* = 0.003) and true HGS (males: *F*_1,405_ = 4.362, *p* = 0.037; females: *F*_1,658_ = 9.928, *p* = 0.002). Post-hoc analysis showed significant differences between pre and post time-points in matched HC for both scores and sexes (all *p* < 0.0000) as well as in female patients for the Δ*HGS* score only (*p* = 0.0005). No significant group differences were observed at any time point. (Fig. 5).

The interaction plots provide valuable insights into the trajectory of true HGS and Δ*HGS* among patients with stroke and MDD in comparison to matched HC for both males and females from pre to post time-points (Fig. 5). For both sexes, true HGS showed a less strong decline from pre to post time-points in patients compared to their matched HC groups. Δ*HGS*, in contrast, demonstrated a significant increase from pre- to post-time-points in all groups, indicating an improvement in HGS relative to the expected HGS predicted based on anthropometrics and age. This increase seems to be stronger in patients compared to controls.

To assess whether the change between pre and post time-points differed significantly between groups, we calculated the difference scores for both true HGS and Δ*HGS* by subtracting the pre time-point values from the post time-point values for each group, separately for males and females. Independent t-tests were then conducted to compare these difference scores between patients and matched HC groups. This approach allows for a direct comparison of the magnitude and direction of change across groups.

In stroke, significant differences were observed only in males who exhibited smaller declines in true HGS compared to matched HC (stroke: -6.80 kg vs matched HC: -11.95 kg, *p* = 0.035) and larger increases in Δ*HGS* (stroke: 3.59 kg vs matched HC: 1.94 kg, *p* = 0.001). MDD patients of both sexes showed significant differences. For true HGS, patients demonstrated smaller decline than matched HC (males: MDD: -6.30 kg vs matched HC: -11.66 kg, *p* = 0.037; females: MDD: -4.50 kg vs matched HC: -9.43 kg, *p* = 0.002). Conversely, for Δ*HGS*, patients exhibited greater increases (males: 3.41 kg vs 2.09 kg, *p* = 0.02; females: 3.17 kg vs 2.26 kg, *p* = 0.003).

## Discussion

In this study, we explored whether HGS can be predicted based on anthropometric and demographic measurements and sought to uncover biological insights by analyzing the predicted HGS values. We developed a new individual-level score Δ*HGS* that captures how HGS deviates from the expected value based on an individual’s characteristics. To this end, we used data from the UK Biobank.

First, we performed ML analysis to predict combined HGS using anthropometric factors (i.e., BMI, height, and WHR) and demographic parameters age and sex. The use of combined HGS, the sum of the grip strengths of both hands, as the target was particularly used as a comprehensive measure of overall strength [33], mitigating the potential biases introduced by handedness or unilateral strength differences [34]. We developed sex-specific models to account for known differences in HGS between males and females, specifically HGS values are consistently higher in males compared to females across all age ranges [15,22–24]. We employed linear SVM and RF regression models for HGS prediction across different sample sizes within a CV scheme. The inclusion of a nonlinear model (RF) alongside the linear SVM was based on the documented nonlinear relationship between age and HGS (Chandrasekaran et al. [20]). In our analysis, linear SVM outperformed RF in predicting combined HGS, achieving higher performance for both sexes. The prediction accuracy improved with larger sample sizes, but smaller samples exhibited higher variance, highlighting the significance of using enough data to adequately capture the variability in demographic and anthropometric factors and their relationship with HGS (Fig. 2, Table 4).

The linear SVM model identified that height and BMI positively contributed to HGS prediction in both sexes, while WHR and age contributed negatively. The magnitude of contributions differed by sex: height and BMI showed a stronger positive contribution in males compared to females (males: FI_height_ = 4.94, FI_BMI_ = 3.00; females: FI_height_ = 3.27, FI_BMI_ = 0.67). WHR had lower contributions in females than males (males: FI_WHR_ = -2.57; females: FI_WHR_ = -0.53). Age contributed negatively for both sexes, reflecting the expected decline of HGS with age (males: -2.83; females: -3.07). These findings align with known associations between these variables and HGS (see the “Model Validation” section). The varying feature importance between sexes taken together with the higher prediction accuracy for males highlight the complex interplay of anthropometric and demographic factors and their association with HGS. The differential contribution of weight-related factors to HGS in females compared to males may be due to sex-specific differences in body composition. Females generally have higher body fat and lower muscle mass than males which could explain why weight-related variables are less predictive of HGS in females [55]. Additionally, hormonal factors, particularly estrogen levels, play a significant role in muscle strength and function in females, potentially overshadowing the influence of weight-related variables on HGS [56].

Our results are in line with the well-established positive relationship between HGS and both BMI and height across various populations and age groups (e.g., M. A. Agtuahene et al. [19] and Yong-Hao Pua et al. [17]). B. Bhattacharjee et al. [18] have reported an inverse relationship between HGS and WHR, which reflects the proportion of abdominal fat relative to hip circumference. Age shows variations in HGS across different age groups. HGS generally declines with age, but the relationship is complex and influenced by sex [16,21]. HGS typically increases during childhood and adolescence, peaks in early adulthood, and then declines with age, particularly after 40 [16]. Vianna et al. [21] found that the onset of HGS decline differs between sexes, beginning earlier in men (around age 30) compared to women (around age 50). The aging process results in progressive muscle loss, impacting HGS. De Araújo et al. [16], identified various factors associated with low HGS in older adults, including age-related differences, emphasizing the multifaceted nature of HGS decline across the lifespan. Given this nonlinear relationship between age and HGS, it is intriguing to consider that our result showed the linear SVM model to be more accurate than the nonlinear RF model. This may be explained by the high mean age in the UKB data. De Araújo et al. [16] also revealed that factors such as socioeconomic status, physical activity levels, and chronic health conditions can influence HGS in older populations, indicating that HGS is also determined by environmental and lifestyle factors. A previous ML study also demonstrated a high prediction accuracy using posture, anthropometric, and demographic variables [57]. This suggests that adding further variables with more detailed body measurements, environmental and lifestyle can help in increasing accuracy. However, large datasets with those variables are currently not available.

The predictions obtained by our models exhibited a bias (Additional file 1: Figure S2), with residuals correlating with the target. To address this, we applied a bias-correction method [40], resulting in bias-free predictions resulting in enhanced accuracy (Fig. 3). The difference between the true HGS and bias-free predicted HGS (Δ*HGS* = *true HGS* –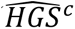) was then calculated as a novel score which was then used for further analysis together with the measured or true HGS.

We explored the neurobiological underpinnings of muscular strength by analyzing the relationship between GMV and two HGS scores. We found that higher HGS is associated with higher GMV in key brain regions involved in motor but also regions playing a role in other functions (Fig. 4). These findings suggest that HGS as a measure of physical capability is reflected in structural brain integrity. However, it is crucial to consider potential confounding effects in these associations, particularly the influence of age. Age has a strong effect on both HGS and GMV and as individuals age, both muscle strength and brain volume typically decrease. Other potential confounders, such as physical activity levels, body composition (e.g., muscle mass, body fat percentage), nutritional status, hormonal factors, genetic predisposition, and socioeconomic status, should also be considered. These factors may independently influence both HGS and GMV, potentially complicating the interpretation of their relationship. Existing literature has shown that HGS is linked to brain structures in frontal, temporal, subcortical, and cerebellar regions. Our findings also revealed associations between HGS and these areas. For instance, Jiang et al. [1] reported widespread positive associations, especially in subcortical regions and temporal cortices, even after controlling for various confounders including age, sex, education level, socioeconomic status, BMI, height, and WHR. Similarly, Meysami et al. [58] reported that greater HGS is associated with larger hippocampal volume, and also stronger dominant HGS was related to larger frontal lobe volumes in older adults. In our results, we found that the hippocampal as well as thalamus regions were associated with HGS in both males and females, along with specific associations in the frontal regions (see Additional file 1: Table S3).

The Δ*HGS* score demonstrated stronger and more widely distributed correlations with GMV compared to true HGS, particularly in motor-related brain regions (Fig. 4). These correlations were significant across various cortical, subcortical, and cerebellar brain regions in both males and females (Fig. 4). The intersection of significant regions between the sexes revealed 667 regions (only 24 for true HGS), while 698 and 755 significant regions were identified in males and females, respectively. Overall, our results suggest that Δ*HGS* is better at capturing the neurobiological basis of muscular strength in both sexes compared to true HGS, suggesting that Δ*HGS* may reflect unique aspects of brain structure beyond what true HGS alone reveals. In contrast to the GMV-true HGS relationships which were all positive, the associations to Δ*HGS* were all negative. The negative GMV-Δ*HGS* correlations suggest that individuals with lower true HGS compared to their predicted HGS tend to have larger GMV. This suggests better preserved brain volume despite lower strength than expected. This relationship becomes more intriguing when considering factors influencing the difference in HGS. Negative Δ*HGS* values might signal an accelerated decline in strength, in which the person is experiencing a more rapid loss of strength than expected for their age or possibly reflecting age-related health concerns that disproportionately affect muscle strength, as muscle strength is a reliable indicator of overall health status in aging populations [7]. Speculatively, a high negative Δ*HGS* could suggest unfavorable body composition or sarcopenic obesity, a condition where muscle loss is combined with excess fat [59].

We then investigated whether changes over time in true HGS and Δ*HGS* differed between patients with either stroke or MDD and their corresponding matched HC in a longitudinal design. We observed a significant reduction in change HGS scores (difference between pre and post disease onset) in both stroke and MDD patients compared to their corresponding matched HC (Table 3), with HC showing a significant decrease in HGS but patients no or less change. Among stroke patients, the decrease in HGS is likely to happen due to damage of the upper motor neuron in the precentral gyrus or of its descending output, i.e., the corticospinal tract, leading to motor impairment, especially of the contralateral hand. Stroke patients typically have lower HGS compared to HC, especially in the early stages post-stroke [60]. ANOVA results revealed sex-specific patterns, with male stroke patients showing significant interactions in both true HGS (*p* = 0.036) and Δ*HGS* (*p* = 0.002), while no significant interactions were observed in female stroke patients. These sex differences align with previous research suggesting different recovery patterns between males and females post-stroke [61]. Importantly, this cannot be because of differing recovery times as the time between disease onset and post assessment was not significantly different between males and females (*p* = 0.59).

Interestingly, despite these longitudinal differences, both HGS and Δ*HGS* were not effective in differentiating between patients and matched HC at the post time-point. This lack of distinction may be attributed to several factors, including the limited number of stroke patients with motor area lesions, potentially masking the effects of these impairments on HGS scores. Importantly, ischemic lesions outside key motor regions or the corticospinal tract do not usually lead to reductions in grip force. In our cohort of 56 stroke patients (1 female patient hemorrhagic), identified using ICD-10 codes without including self-reported cases, post-stroke neuroimaging data were available for 25 patients (44.64%). Among these, 14 patients (56%) presented lesions in motor-related regions, with 9 of them exhibiting lesions affecting the contralateral hand. Additional factors include the substantial time elapsed between pre and post assessments in both disease cohorts which may reflect the effects of treatment and rehabilitation efforts over time. Specifically, in stroke patients, the average interval between pre and post assessments was approximately 9.26 years for males and 9.5 years for females, with post-stroke assessments occurring between 0.18-11.14 years for males and 0.04-9.78 years for females after stroke onset.

In MDD patients, the reduction in HGS may be due to the multifactorial impact of MDD on physical health [62], which is attributed to the complex interplay between depression and physical health. Ganipineni et al. [10] found that individuals with depressive symptoms tend to have lower HGS compared to those without. Trivedi [62] showed that MDD not only affects mental health but also has wide-ranging effects on physical health through various mechanisms. These include decreased physical activity, changes in appetite and nutrition, sleep disturbances, hormonal and inflammatory alterations, and potential medication side effects, all of which can contribute to reduced grip strength. ANOVA results showed significant interactions in true HGS (males: *p* = 0.037; females: *p* = 0.002) and Δ*HGS* (males: *p* = 0.021; females: *p* = 0.003) for both sexes, in contrast to the sex-specific patterns observed in stroke patients. However, despite these longitudinal differences, both HGS and Δ*HGS* were ineffective in distinguishing MDD patients from matched HC at the post time-point. This finding may reflect the prolonged time elapsed between pre and post assessments, with the mean time intervals of 9.46 years for males and 9.3 years for females. Post-MDD assessments occurred within 0.02-9.63 years for males and 0.02-11.98 years for females, highlighting the influence of ongoing treatment and rehabilitation. Further research could provide deeper insights into the relationship between muscle function and diseases, supporting early detection, targeted prevention, and monitoring of disease progression in neurological and psychiatric conditions.

Our study faced several limitations that should be addressed in future research. Firstly, the demographic (age and sex) and anthropometric variables (BMI, height, and WHR) may not capture all relevant factors influencing muscle strength, potentially overlooking other predictors like more detailed body measurements, genetics, and lifestyle factors. Secondly, the study sample was drawn from the UKB, which may not be representative of other populations, potentially limiting the generalizability of the findings to more diverse demographic groups. Thirdly, the study used a limited number of ML models, focusing on linear SVM and RF, which may not fully capture the complexity of the data. Although we covered linear and non-linear models, exploring other models could potentially improve prediction accuracy. Lastly, the longitudinal study was constrained by small sample sizes for stroke and MDD patients, which could affect the statistical power and reliability of the results. Here, we relied on incidental data from the UKB, which comes with limitations such as low sample sizes, the small number of stroke patients with motor area related lesions, and lack of detailed information regarding medication and rehabilitation.

## Conclusion

This study demonstrates that HGS can be predicted using demographic and anthropometric variables at moderate accuracy through machine learning models, particularly linear SVM. The novel Δ*HGS* score, representing the difference between true and bias-free predicted HGS, showed stronger and widely distributed correlations with GMV compared to true HGS, especially in motor-related brain regions. This suggests Δ*HGS* maybe a more sensitive biomarker for brain health assessment. Both true HGS and Δ*HGS* did not capture longitudinal differences between patients and matched HC. Further research is needed to validate these results in more diverse populations and explore the mechanisms linking HGS changes to specific diseases. Overall, this study provides a foundation for enhancing the utility of HGS as a biomarker in neurological and psychiatric research and clinical practice.

## Data Availability

All data used in this study are publicly available through the UK Biobank, accessible via their standard data access procedure at http://www.ukbiobank.ac.uk/.

## Abbreviations

HGS: Handgrip strength
HC: Healthy controls
MDD: Major depressive disorder
GMV: Grey matter volume
ML: Machine learning
SVM: Support Vector Machine
RF: Random Forest
FI: Feature importance
MAE: Mean absolute error
CCC: Concordance correlation coefficient

## Supplementary Information

The online version contains supplementary material available at the additional supplementary file (supplementary.pdf)

**Additional file 1: Table S1.** Summary of the characteristics of the HC participants for matched sample analysis. **Table S2.** Summary of the characteristics of the non-imaging HC participants on test dataset for reassessment reliability analysis. **Table S3.** Summary of the top 10 subcortical regions with strongest correlation with true HGS. **Table S4.** Summary of the top 10 subcortical regions with strongest correlation with Δ*HGS*. **Figure S1.** Flowchart of UKB participants selection and included in the analysis. **Figure S2.** Scatter plots depicting the relationship between various scores of 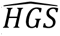 and 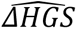 without bias-correction versus true HGS on the independent non-brain-imaging HC test dataset, for males and females.

## Acknowledgments

This research has been conducted using data from UK Biobank resources (application number 41655). All data used in this study are publicly accessible from UK Biobank via their standard data access procedure (http://www.ukbiobank.ac.uk/).

## Authors’ contributions

All authors read and approved the final manuscript. We use the CRediT contributor role taxonomy to describe individual contributions to the paper. Conceptualization: K.N., S.B.E, C.G., K.R.P, L.H., V.M.; Data curation: K.N., C.T, V.K., K.R.P.; Formal analysis: K.N.; Methodology: K.N., S.B.E., L.H, V.M, C.G., K.R.P; Supervision: C.G., S.B.E., K.R.P.; Visualization: K.N., Writing—original draft: K.N.; Writing—review & editing: K.N., S.B.E, G. A, L.H., C.T., V.K, F.R., V.M., C.G., K.R.P.

## Declarations

### Ethics approval and consent to participate

The UK Biobank study was approved by the North West Multicenter Research Ethics Committee (No. 16/NW/0274), with written informed consent obtained from all participants. A re-analysis of the anonymized data was approved by the ethics committee of the Heinrich Heine University Düsseldorf (2018-317-RetroDEuA).

### Consent for publication

Not applicable.

## Reference

[1] Jiang R, Westwater ML, Noble S, Rosenblatt M, Dai W, Qi S, et al. Associations between grip strength, brain structure, and mental health in > 40,000 participants from the UK Biobank. BMC Medicine 2022;20:286. 10.1186/s12916-022-02490-2.

[2] Roberts HC, Denison HJ, Martin HJ, Patel HP, Syddall H, Cooper C, et al. A review of the measurement of grip strength in clinical and epidemiological studies: towards a standardised approach. Age and Ageing 2011;40:423*–*9. 10.1093/ageing/afr051.

[3] Duchowny KA, Ackley SF, Brenowitz WD, Wang J, Zimmerman SC, Caunca MR, et al. Associations Between Handgrip Strength and Dementia Risk, Cognition, and Neuroimaging Outcomes in the UK Biobank Cohort Study. JAMA Netw Open 2022;5:e2218314. 10.1001/jamanetworkopen.2022.18314.

[4] Celis-Morales CA, Petermann F, Hui L, Lyall DM, Iliodromiti S, McLaren J, et al. Associations Between Diabetes and Both Cardiovascular Disease and All-Cause Mortality Are Modified by Grip Strength: Evidence From UK Biobank, a Prospective Population-Based Cohort Study. Diabetes Care 2017;40:1710*–*8. 10.2337/dc17-0921.

[5] Yates T, Zaccardi F, Dhalwani NN, Davies MJ, Bakrania K, Celis-Morales CA, et al. Association of walking pace and handgrip strength with all-cause, cardiovascular, and cancer mortality: a UK Biobank observational study. European Heart Journal 2017;38:3232*–*40. 10.1093/eurheartj/ehx449.

[6] Soltanisarvestani M, Lynskey N, Gray S, Gill JMR, Pell JP, Sattar N, et al. Associations of grip strength and walking pace with mortality in stroke survivors: A prospective study from UK Biobank. Scandinavian Journal of Medicine & Science in Sports 2023;33:1190*–*200. 10.1111/sms.14352.

[7] Nishita Y, Nakamura A, Kato T, Otsuka R, Iwata K, Tange C, et al. Links Between Physical Frailty and Regional Gray Matter Volumes in Older Adults: A Voxel-Based Morphometry Study. J Am Med Dir Assoc 2019;20:1587–1592.e7. 10.1016/j.jamda.2019.09.001.

[8] Vaishya R, Misra A, Vaish A, Ursino N, D*’*Ambrosi R. Hand grip strength as a proposed new vital sign of health: a narrative review of evidences. Journal of Health, Population and Nutrition 2024;43:7. 10.1186/s41043-024-00500-y.

[9] Zheng H, He Q, Xu H, Zheng X, Gu Y. Lower grip strength and insufficient physical activity can increase depressive symptoms among middle-aged and older European adults: a longitudinal study. BMC Geriatrics 2022;22:696. 10.1186/s12877-022-03392-x.

[10] Ganipineni VDP, Idavalapati ASKK, Tamalapakula SS, Moparthi V, Potru M, Owolabi OJ. Depression and Hand-Grip: Unraveling the Association. Cureus n.d.;15:e38632. 10.7759/cureus.38632.

[11] Zaccagni L, Toselli S, Bramanti B, Gualdi-Russo E, Mongillo J, Rinaldo N. Handgrip Strength in Young Adults: Association with Anthropometric Variables and Laterality. Int J Environ Res Public Health 2020;17:4273. 10.3390/ijerph17124273.

[12] Tscherpel C, Dern S, Hensel L, Ziemann U, Fink GR, Grefkes C. Brain responsivity provides an individual readout for motor recovery after stroke. Brain 2020;143:1873. 10.1093/brain/awaa127.

[13] Bonkhoff AK, Rehme AK, Hensel L, Tscherpel C, Volz LJ, Espinoza FA, et al. Dynamic connectivity predicts acute motor impairment and recovery post-stroke. Brain Commun 2021;3:fcab227. 10.1093/braincomms/fcab227.

[14] McGrath R, Johnson N, Klawitter L, Mahoney S, Trautman K, Carlson C, et al. What are the association patterns between handgrip strength and adverse health conditions? A topical review. SAGE Open Medicine 2020;8:2050312120910358. 10.1177/2050312120910358.

[15] Wunderle V, Kuzu TD, Tscherpel C, Fink GR, Grefkes C, Weiss PH. Age- and sex-related changes in motor functions: a comprehensive assessment and component analysis. Front Aging Neurosci 2024;16. 10.3389/fnagi.2024.1368052.

[16] de Araújo Amaral C, Amaral TLM, Monteiro GTR, de Vasconcellos MTL, Portela MC. Factors associated with low handgrip strength in older people: data of the Study of Chronic Diseases (Edoc-I). BMC Public Health 2020;20:395. 10.1186/s12889-020-08504-z.

[17] Pua Y-H, Tay L, Clark RA, Thumboo J, Tay E-L, Mah S-M, et al. Associations of height, weight, and body mass index with handgrip strength: A Bayesian comparison in older adults. Clinical Nutrition ESPEN 2023;54:206*–*10. 10.1016/j.clnesp.2023.01.028.

[18] Bhattacharjee B, Ghosh J, Bhattacharjee A, Singh K, Roychowdhury S, Roy A, et al. Association of handgrip strength with blood pressure, waist hip ratio, visceral adiposity index, C-reactive protein among adult population of Kolkata: A hospital based cross-sectional observational study. Clinical Nutrition ESPEN 2023;58:523. 10.1016/j.clnesp.2023.09.291.

[19] Agtuahene MA, Quartey J, Kwakye S. Influence of hand dominance, gender, and body mass index on hand grip strength. S Afr J Physiother 2023;79:1923. 10.4102/sajp.v79i1.1923.

[20] Chandrasekaran B, Ghosh A, Prasad C, Krishnan K, Chandrasharma B. Age and Anthropometric Traits Predict Handgrip Strength in Healthy Normals. J Hand Microsurg 2016;02:58*–*61. 10.1007/s12593-010-0015-6.

[21] Vianna L, Oliveira R, Araujo CG. Age-Related Decline in Handgrip Strength Differs According to Gender. Journal of Strength and Conditioning Research / National Strength & Conditioning Association 2007;21:1310*–*4. 10.1519/R-23156.1.

[22] Kim J, Kim Y, Oh JW, Lee S. Sex differences of the association between handgrip strength and health-related quality of life among patients with cancer. Sci Rep 2024;14:9876. 10.1038/s41598-024-60710-6.

[23] Komeyer V, Eickhoff SB, Grefkes C, Patil KR, Raimondo F. A framework for confounder considerations in AI-driven precision medicine 2024:2024.02.02.24302198. 10.1101/2024.02.02.24302198.

[24] Spruit MA, Sillen MJH, Groenen MTJ, Wouters EFM, Franssen FME. New normative values for handgrip strength: results from the UK Biobank. J Am Med Dir Assoc 2013;14:775.e5–11. 10.1016/j.jamda.2013.06.013.

[25] Rehme AK, Fink GR, von Cramon DY, Grefkes C. The Role of the Contralesional Motor Cortex for Motor Recovery in the Early Days after Stroke Assessed with Longitudinal fMRI. Cerebral Cortex 2011;21:756*–*68. 10.1093/cercor/bhq140.

[26] Hwang J, Lee J, Lee K-S. A deep learning-based method for grip strength prediction: Comparison of multilayer perceptron and polynomial regression approaches. PLoS One 2021;16:e0246870. 10.1371/journal.pone.0246870.

[27] Allen N, Sudlow C, Downey P, Peakman T, Danesh J, Elliott P, et al. UK Biobank: Current status and what it means for epidemiology. Health Policy and Technology 2012;1:123*–*6. 10.1016/j.hlpt.2012.07.003.

28. [28] Collins R. UK Biobank: protocol for a large-scale prospective epidemiological resource. 2007. http://www.ukbiobank.ac.uk/wp-content/uploads/2011/11/UKBiobank-Protocol.pdf (21 March 2017). n.d.

[29] UK Biobank. Algorithmicallydefined outcomes(ADOs) 2022. https://biobank.ndph.ox.ac.uk/showcase/refer.cgi?id=460 (accessed June 20, 2024).

[30] UK Biobank. First Occurrence of Health Outcomes 2019. https://biobank.ndph.ox.ac.uk/showcase/refer.cgi?id=593 (accessed August 1, 2024).

[31] Glanville KP, Coleman JRI, Howard DM, Pain O, Hanscombe KB, Jermy B, et al. Multiple measures of depression to enhance validity of major depressive disorder in the UK Biobank. BJPsych Open 2021;7:e44. 10.1192/bjo.2020.145.

[32] Firth J, Stubbs B, Vancampfort D, Firth JA, Large M, Rosenbaum S, et al. Grip Strength Is Associated With Cognitive Performance in Schizophrenia and the General Population: A UK Biobank Study of 476559 Participants. Schizophr Bull 2018;44:728*–*36. 10.1093/schbul/sby034.

[33] Bobos P, Nazari G, Lu Z, MacDermid JC. Measurement Properties of the Hand Grip Strength Assessment: A Systematic Review With Meta-analysis. Arch Phys Med Rehabil 2020;101:553*–*65. 10.1016/j.apmr.2019.10.183.

[34] Petersen P, Petrick M, Connor H, Conklin D. Grip Strength and Hand Dominance: Challenging the 10% Rule. The American Journal of Occupational Therapy 1989;43:444*–*7. 10.5014/ajot.43.7.444.

[35] Gómez-Campos R, Vidal Espinoza R, de Arruda M, Ronque ERV, Urra-Albornoz C, Minango JC, et al. Relationship between age and handgrip strength: Proposal of reference values from infancy to senescence. Front Public Health 2023;10. 10.3389/fpubh.2022.1072684.

[36] Hamdan S, More S, Sasse L, Komeyer V, Patil KR, Raimondo F. Julearn: an easy-to-use library for leakage-free evaluation and inspection of ML models 2023. 10.48550/arXiv.2310.12568.

[37] Pedregosa F, Varoquaux G, Gramfort A, Michel V, Thirion B, Grisel O, et al. Scikit-learn: Machine Learning in Python. Journal of Machine Learning Research 2011;12:2825*–*30.

[38] R: Fast Heuristics For The Estimation Of the C Constant Of A… n.d. https://search.r-project.org/CRAN/refmans/LiblineaR/html/heuristicC.html (accessed June 9, 2024).

[39] Young DS. Handbook of Regression Methods. New York: Chapman and Hall/CRC; 2017. 10.1201/9781315154701.

[40] Beheshti I, Nugent S, Potvin O, Duchesne S. Bias-adjustment in neuroimaging-based brain age frameworks: A robust scheme. NeuroImage: Clinical 2019;24:102063. 10.1016/j.nicl.2019.102063.

[41] Lin LI. A concordance correlation coefficient to evaluate reproducibility. Biometrics 1989;45:255*–*68.

[42] Sudlow C, Gallacher J, Allen N, Beral V, Burton P, Danesh J, et al. UK biobank: an open access resource for identifying the causes of a wide range of complex diseases of middle and old age. PLoS Med 2015;12:e1001779. 10.1371/journal.pmed.1001779.

[43] Miller KL, Alfaro-Almagro F, Bangerter NK, Thomas DL, Yacoub E, Xu J, et al. Multimodal population brain imaging in the UK Biobank prospective epidemiological study. Nat Neurosci 2016;19:1523*–*36. 10.1038/nn.4393.

[44] Alfaro-Almagro F, Jenkinson M, Bangerter NK, Andersson JLR, Griffanti L, Douaud G, et al. Image processing and Quality Control for the first 10,000 brain imaging datasets from UK Biobank. NeuroImage 2018;166:400*–*24. 10.1016/j.neuroimage.2017.10.034.

[45] Halchenko YO, Meyer K, Poldrack B, Solanky DS, Wagner AS, Gors J, et al. DataLad: distributed system for joint management of code, data, and their relationship. Journal of Open Source Software 2021;6:3262. 10.21105/joss.03262.

[46] Wagner AS, Waite LK, Wierzba M, Hoffstaedter F, Waite AQ, Poldrack B, et al. FAIRly big: A framework for computationally reproducible processing of large-scale data. Sci Data 2022;9:80. 10.1038/s41597-022-01163-2.

[47] Schaefer A, Kong R, Gordon EM, Laumann TO, Zuo X-N, Holmes AJ, et al. Local-Global Parcellation of the Human Cerebral Cortex from Intrinsic Functional Connectivity MRI. Cereb Cortex 2018;28:3095*–*114. 10.1093/cercor/bhx179.

[48] Tian Y, Margulies DS, Breakspear M, Zalesky A. Topographic organization of the human subcortex unveiled with functional connectivity gradients. Nat Neurosci 2020;23:1421*–*32. 10.1038/s41593-020-00711-6.

[49] Diedrichsen J, Balsters JH, Flavell J, Cussans E, Ramnani N. A probabilistic MR atlas of the human cerebellum. NeuroImage 2009;46:39*–*46. 10.1016/j.neuroimage.2009.01.045.

[50] Zhao Q-Y, Luo J-C, Su Y, Zhang Y-J, Tu G-W, Luo Z. Propensity score matching with R: conventional methods and new features. Ann Transl Med 2021;9:812. 10.21037/atm-20-3998.

[51] Salkind NJ. Encyclopedia of Research Design. SAGE; 2010.

[52] Glass GV, Peckham PD, Sanders JR. Consequences of Failure to Meet Assumptions Underlying the Fixed Effects Analyses of Variance and Covariance. Review of Educational Research 1972;42:237*–*88. 10.3102/00346543042003237.

[53] Harwell MR, Rubinstein EN, Hayes WS, Olds CC. Summarizing Monte Carlo Results in Methodological Research: The One- and Two-Factor Fixed Effects ANOVA Cases. Journal of Educational Statistics 1992;17:315*–*39. 10.3102/10769986017004315.

[54] Lumley T, Diehr P, Emerson S, Chen L. The importance of the normality assumption in large public health data sets. Annu Rev Public Health 2002;23:151*–*69. 10.1146/annurev.publhealth.23.100901.140546.

[55] Liao K-H. Hand Grip Strength in Low, Medium, and High Body Mass Index Males and Females. Middle East J Rehabil Health Stud 2016;3. 10.17795/mejrh-33860.

[56] Arvandi M, Strasser B, Meisinger C, Volaklis K, Gothe RM, Siebert U, et al. Gender differences in the association between grip strength and mortality in older adults: results from the KORA-age study. BMC Geriatrics 2016;16:201. 10.1186/s12877-016-0381-4.

[57] Hwang J, Lee J, Lee K-S. A deep learning-based method for grip strength prediction: Comparison of multilayer perceptron and polynomial regression approaches. PLoS ONE 2021;16:e0246870. 10.1371/journal.pone.0246870.

[58] Meysami S, Raji CA, Glatt RM, Popa ES, Ganapathi AS, Bookheimer T, et al. Handgrip Strength Is Related to Hippocampal and Lobar Brain Volumes in a Cohort of Cognitively Impaired Older Adults with Confirmed Amyloid Burden. J Alzheimers Dis n.d.;91:999*–*1006. 10.3233/JAD-220886.

[59] Stenholm S, Harris TB, Rantanen T, Visser M, Kritchevsky SB, Ferrucci L. Sarcopenic obesity - definition, etiology and consequences. Curr Opin Clin Nutr Metab Care 2008;11:693*–*700. 10.1097/MCO.0b013e328312c37d.

[60] Stock R, Thrane G, Askim T, Anke A, Mork PJ. Development of grip strength during the first year after stroke. J Rehabil Med 2019;51:248*–*56. 10.2340/16501977-2530.

[61] Poggesi A, Insalata G, Papi G, Rinnoci V, Donnini I, Martini M, et al. Gender differences in post-stroke functional outcome at discharge from an intensive rehabilitation hospital. European Journal of Neurology 2021;28:1601*–*8. 10.1111/ene.14769.

[62] Trivedi MH. The Link Between Depression and Physical Symptoms. Prim Care Companion J Clin Psychiatry 2004;6:12*–*6.

