## Supplementary Information for "Machine Learning-Driven Correction of Handgrip Strength: A Novel Biomarker for Neurological and Health Outcomes in the UK Biobank"

**Table S1:** Summary of the characteristics of the HC participants for matched sample analysis.

| Population | HC with brain-imaging | | | |
|---|---|---|---|---|
| Data set | Baseline assessment visit | First repeat assessment visit | Imaging visit | First repeat imaging visit |
| Both gender | | | | |
| Number | 25,632 | 3,897 | 23,926 | 2,512 |
| Age, mean (SD) | 54.77 (7.55) | 59.85 (7.35) | 64.21 (7.74) | 64.4 (7.2) |
| BMI, mean (SD) | 26.22 (4.1) | 26.21 (4.13) | 26.18 (4.31) | 26.01 (4.3) |
| Height, mean (SD) | 169.65 (9.14) | 169.24 (9.2) | 169.01 (9.17) | 169.27 (9.23) |
| WHR, mean (SD) | 0.85 (0.09) | 0.87 (0.08) | 0.87 (0.09) | 0.88 (0.09) |
| Combined HGS, mean (SD) | 65.84 (21.32) | 54.77 (20.35) | 60.00 (20.21) | 57.53 (20.56) |
| Right dominant hand | 90.70% | 91.45% | 91.11% | 90.57% |
| Female | | | | |
| Number | 13,714 | 2,013 | 12,849 | 1,359 |
| Age, mean (SD) | 54.05 (7.35) | 58.98 (7.21) | 63.48 (7.55) | 63.71 (6.94) |
| BMI, mean (SD) | 25.69 (4.39) | 25.7 (4.42) | 25.77 (4.64) | 25.57 (4.6) |
| Height, mean (SD) | 163.5 (6.13) | 162.7 (6.04) | 162.9 (6.26) | 163.13 (6.23) |
| WHR, mean (SD) | 0.8 (0.07) | 0.81 (0.06) | 0.82 (0.07) | 0.83 (0.07) |
| Combined HGS, mean (SD) | 50.83 (11.68) | 40.32 (11.5) | 46.53 (11.37) | 44.24 (12.03) |
| Right dominant hand | 92.21% | 93.59% | 92.72% | 92.79% |
| Male | | | | |
| Number | 11,918 | 1,884 | 11,077 | 1,153 |
| Age, mean (SD) | 55.59(7.69) | 60.77 (7.38) | 65.06 (7.87) | 65.21 (7.4) |
| BMI, mean (SD) | 26.82 (3.65) | 26.75 (3.73) | 26.67 (3.83) | 26.52 (3.85) |
| Height, mean (SD) | 176.72 (6.54) | 176.22 (6.46) | 176.1 (6.54) | 176.51 (6.57) |
| WHR, mean (SD) | 0.92 (0.06) | 0.92 (0.06) | 0.93 (0.06) | 0.94 (0.06) |
| Combined HGS, mean (SD) | 83.11 (16.23) | 70.2 (15.92) | 75.61 (16.66) | 73.20 (17.22) |
| Right dominant hand (%) | 88.97% | 89.17% | 89.24% | 87.94% |

**Table S2:** Summary of the characteristics of the non-imaging HC participants on test dataset for reassessment reliability analysis.

| Population | HC non-brain-imaging | |
|---|---|---|
| Data set | Baseline assessment visit | First repeat assessment visit |
| Female | | |
| Number | 162 | 162 |
| Age, mean (SD) | 55.92 (7.32) | 60.26 (7.38) |
| BMI, mean (SD) | 25.73 (4.01) | 25.82 (4.12) |
| Height, mean (SD) | 162.78 (6.08) | 162.2 (6.01) |
| WHR, mean (SD) | 0.8 (0.07) | 0.82 (0.06) |
| Combined HGS, mean (SD) | 50.17 (11.09) | 37.24 (11.82) |
| Right dominant hand | 93.21% | 93.21% |
| Male | | |
| Number | 134 | 134 |
| Age, mean (SD) | 56.87(7.35) | 61.31 (7.41) |
| BMI, mean (SD) | 27.22 (3.76) | 27.41 (3.87) |
| Height, mean (SD) | 175.2 (6.29) | 174.74 (6.19) |
| WHR, mean (SD) | 0.92 (0.05) | 0.93 (0.06) |
| Combined HGS, mean (SD) | 81.79 (18.39) | 64.98 (17.85) |
| Right dominant hand (%) | 91.04% | 91.04% |

**Table S3:** Summary of the top 10 subcortical regions with strongest correlation with true HGS.

| Both gender | | Male | | Female | |
|---|---|---|---|---|---|
| regions | $r$ values | regions | $r$ values | regions | $r$ values |
| THA-VAia-lh | 0.119456 | HIP-body-rh | 0.176761 | THA-VAia-lh | 0.113411 |
| | | HIP-body-lh | 0.166598 | | |
| | | mAMY-lh | 0.162900 | | |
| | | mAMY-rh | 0.157426 | | |
| | | HIP-head-l-rh | 0.145028 | | |
| | | HIP-head-l-lh | 0.138328 | | |
| | | lAMY-lh | 0.137043 | | |
| | | THA-DP-rh | 0.133825 | | |
| | | lAMY-rh | 0.129584 | | |
| | | NAc-shell-lh | 0.128321 | | |

**Table S4:** Summary of the top 10 subcortical regions with strongest correlation with $\Delta HGS$.

| Both gender | | Male | | Female | |
|---|---|---|---|---|---|
| regions | $r$ values | regions | $r$ values | regions | $r$ values |
| HIP-body-lh | -0.235108 | HIP-body-rh | -0.256797 | THA-DP-rh | -0.231279 |
| HIP-body-rh | -0.226771 | HIP-body-lh | -0.256456 | THA-DP-lh | -0.225983 |
| mAMY-rh | -0.221798 | mAMY-rh | -0.225924 | mAMY-rh | -0.217673 |
| THA-DP-rh | -0.217565 | HIP-head-l-rh | -0.219348 | THA-VAia-lh | -0.217030 |
| THA-DP-lh | -0.216335 | mAMY-lh | -0.216216 | mAMY-lh | -0.215104 |
| mAMY-lh | -0.215660 | HIP-head-l-lh | -0.214152 | HIP-body-lh | -0.213760 |
| THA-VAia-lh | -0.209914 | THA-VAia-rh | -0.211763 | NAc-shell-lh | -0.198184 |
| THA-VAia-rh | -0.201281 | THA-DP-lh | -0.206688 | THA-VAip-lh | -0.197668 |
| HIP-head-l-rh | -0.196375 | THA-DP-rh | -0.203851 | HIP-body-rh | -0.196745 |
| HIP-head-l-lh | -0.196261 | THA-VAia-lh | -0.202797 | lAMY-lh | -0.231279 |

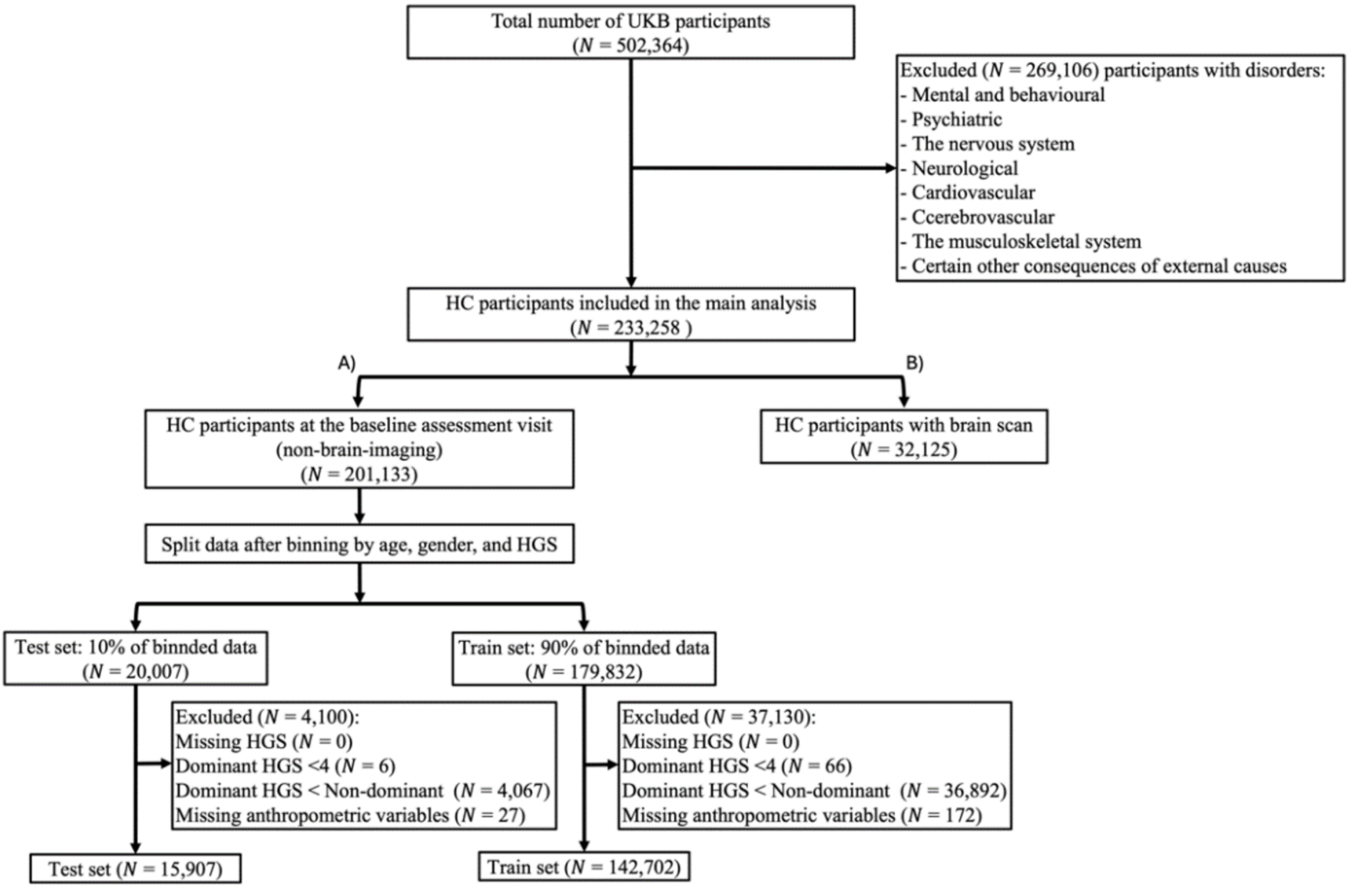


**Figure S1:** Flowchart of UKB participants selection and included in the analysis. The HC population for this study was divided into two distinct sub-datasets, ensuring no overlap. This was achieved by excluding participants who reported different diseases from the total UK Biobank (UKB) participants pool (N = 502,364. A) The first sub-dataset comprises HC participants who did not undergo MRI scans (N = 201,133, 54.68% female, age at baseline = 55.44 ± 8.14 years) and was used for creating the training and test datasets. B) The second sub-dataset includes HC participants who attended imaging assessment visit sessions (N = 32,125, 51.7% female, age at baseline = 54.01 ± 7.35 years). After applying additional exclusion criteria to remove cases with missing data or relevant HGS dominant conditions, the non-brain-imaging data resulted in two distinct datasets: a training dataset (N = 142,702, 56.68% female) and a test dataset (N = 15,907, 56.38% female).

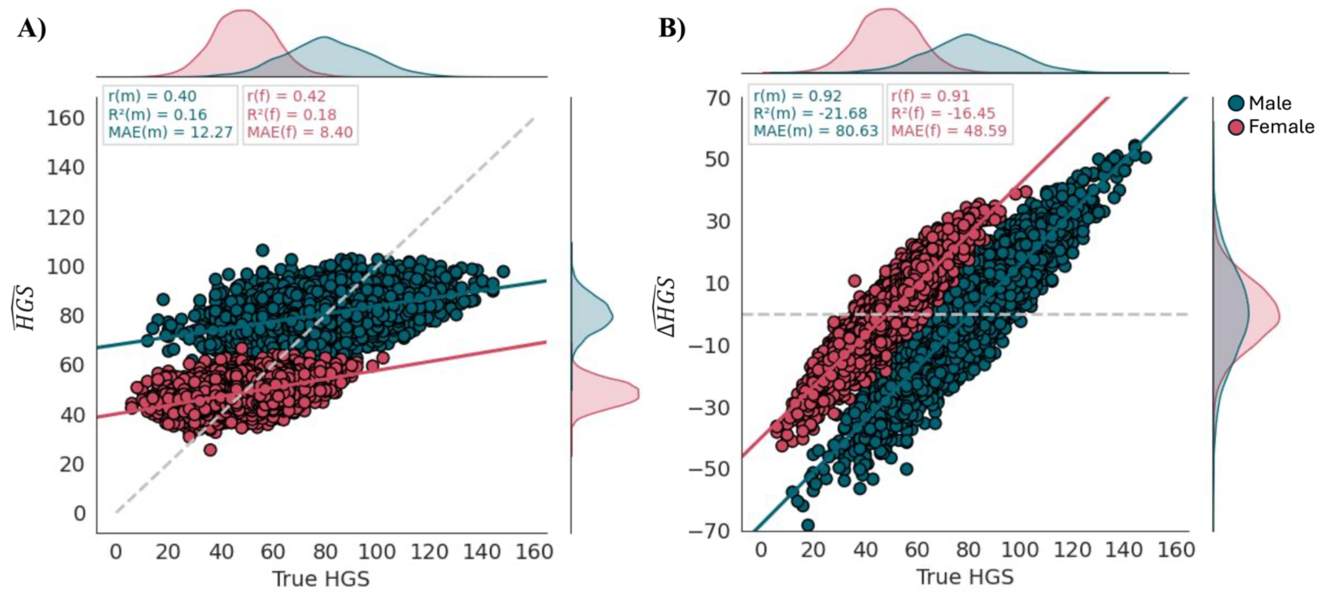


**Figure S2:** Relationship between $\widehat{HGS}$ (A) and $\Delta\widehat{HGS}$ (B) scores versus true HGS on the independent non-brain-imaging HC test dataset, for males (N = 6,938) and females (N = 8,969). **A)** Scatter plot of $\widehat{HGS}$ and true HGS, without bias-correction: for males ($r$ = 0.4, $R^2$ = 0.16, $MAE$ = 12.27) and for females ($r$ = 0.42, $R^2$ = 0.18, $MAE$ = 8.4). **B)** Scatter plot of $\Delta\widehat{HGS}$ and true HGS without the bias-correction: for males ($r$ = 0.92, $R^2$ = -21.68, $MAE$ = 80.63) and for females ($r$ = 0.91, $R^2$ = -16.45, $MAE$ = 48.59). The dashed grey line on the A indicated the identity line (y = x), while the dashed grey line on the B indicated the reference line (y = 0).
